# Integrative Multi-Omic Profiling of cfDNA Methylation and EV-miRNAs Identifies Immunotherapy-Outcome Molecular Subtypes in NSCLC

**DOI:** 10.1101/2025.08.19.25333961

**Authors:** Juan Luis Onieva, Elisabeth Pérez-Ruiz, Laura C Figueroa-Ortiz, José Miguel Jurado, Beatriz Martínez-Gálvez, Jose Carlos Benitez, Antonio Rueda-Domínguez, Isabel Barragán

**Author notes:** Contributed equally (Antonio Rueda-Domínguez, Isabel Barragán).

## Abstract

Non-small cell lung cancer (NSCLC) patients exhibit heterogeneous responses to immunotherapy (IT) with high resistance rates, highlighting the need for precise biomarkers predictive of treatment outcomes. In a prospective cohort study, we longitudinally assessed liquid biopsy samples from NSCLC patients undergoing IT at four distinct time points (T1 pre-treatment, T2 post-second cycle, T3 six months, and T4 one year). We profiled plasma-derived cell-free DNA methylation and extracellular vesicle-associated microRNAs from 79 metastatic NSCLC patients treated with immune checkpoint inhibitors (ICIs). High-dimensional omics data were integrated using Multi-Omics Factor Analysis (MOFA2) to uncover latent molecular subtypes, which we termed MOFA-Derived Clusters (MDCs), independently established at baseline (MDC-T1) and post-second cycle (MDC-T2). Differential expression and methylation analyses, pathway enrichment, and immune phenotyping via flow cytometry were used to characterize the molecular and immunological landscape of each MDC. External validation was performed using independent NSCLC cohorts for miRNAs (Genova et al., 2024, n=54) and methylation (SMC-Cohort, GSE119144, n=57). MDCs captured divergent survival outcomes and reflected biologically coherent processes including angiogenesis, cytoskeletal remodeling, and immune signaling. Projection of MDCs onto later time points (T3, T4) supported the temporal relevance of early molecular signatures. MDCs also displayed immunological correlates via circulating immune cell subsets. Importantly, MDC classifiers demonstrated consistent survival stratification in external cohorts, particularly MDC-T2. This study defines a multi-omic, liquid biopsy–based framework for molecular subtyping in NSCLC to manage ICI treatment. Our MDC signatures reveal clinically meaningful, treatment-informative biology and offer a path toward minimally invasive patient stratification in immuno-oncology.

## Introduction

Lung cancer remains the most frequently diagnosed malignancy and the leading cause of cancer-related mortality worldwide ^1^. Non-small cell lung cancer (NSCLC) accounts for approximately 85% of all lung cancer cases and is primarily composed of two major histological subtypes: lung adenocarcinoma (LUAD) and lung squamous cell carcinoma (LUSC) ^2^. Over the past decade, significant advances in immunotherapy have translated into a marked improvement in overall survival (OS) among patients with NSCLC at the population level ^3^. In particular, immune checkpoint inhibitors (ICIs) have demonstrated clinical benefits in patients with advanced NSCLC^4^, markedly transforming treatment strategies ^5^.

Despite the increasing role of ICIs in NSCLC, a substantial number of patients do not benefit from these therapies, ranging from approximately 20% to 45% ^4,6^. Moreover, immune-related adverse events (irAEs) occur in a subset of patients leading to treatment discontinuation, long-term morbidity, or even fatal outcomes ^7^. These limitations highlight the pressing need for predictive biomarkers with high analytical and clinical validity, capable of accurately stratifying NSCLC patients according to their probability of response to ICIs.

Currently, PD-L1 expression is regarded as the most widely implemented biomarker approved by both the U.S. Food and Drug Administration (FDA) ^8^ and the National Comprehensive Cancer Network (NCCN) ^9^ to guide treatment decisions in patients with metastatic NSCLC. Despite its routine use in clinical practice, the predictive performance of PD-L1 is limited. Expression levels are subject to both inter-and intra-tumoral heterogeneity, and variability is further compounded by discrepancies among commercially available immunohistochemistry assays ^10^. These factors collectively reduce the reliability of PD-L1 as a stand-alone predictive biomarker. In parallel, tumor mutational burden (TMB)—defined as the total number of somatic mutations per megabase in coding regions of tumor DNA—has been approved by the FDA as a pan-cancer biomarker for ICIs therapy, based on findings from the phase II KEYNOTE-158 trial ^11^. TMB is thought to reflect the potential for neoantigen generation, which may enhance tumor immunogenicity. However, despite their theoretical rationale, neither PD-L1 nor TMB has demonstrated sufficient predictive precision to consistently identify patients who will derive durable benefit from ICI treatment. Moreover, both biomarkers require analysis of solid tumor tissue, necessitating invasive biopsy procedures that may not be feasible in all clinical scenarios, thereby limiting their broader applicability in routine care.

In this context, liquid biopsy has emerged as a promising, minimally invasive approach capable of capturing a broad spectrum of tumor-derived components—including circulating tumor cells (CTCs), cell free DNA (cfDNA), and extracellular vesicles (EVs)—from various body fluids such as blood, saliva, and pleural effusion ^12^. Compared to conventional tissue biopsy, liquid biopsy offers several critical advantages: it enables real-time and repeated sampling and facilitates dynamic monitoring of treatment response ^13^. In our study we have focused on the epigenetic landscape of circulating cfDNA and microRNAs (miRNAs) encapsulated in EVs, two biomarkers omics with regulatory functions in cancer biology ^14^. cfDNA, released into the bloodstream primarily through apoptosis and necrosis, reflects the genetic and epigenetic landscape of the tumor. Meanwhile, EV-derived miRNAs serve as an active modulators of intercellular communication, influencing processes such as immune regulation, angiogenesis, and metastatic progression. DNA methylation is one of several epigenetic mechanisms that cells use to control gene expression, in part by promoting or inhibiting the binding of transcription factors to DNA. Conversely, at the post-transcriptional level, microRNAs regulate gene expression by hybridizing to target mRNAs, thereby modulating their translational efficiency or stability. Given these attributes, both cfDNA and EV-associated miRNAs represent promising biomarker candidates for improving patient stratification and guiding precision immunotherapy in NSCLC. Combined with the advantages of liquid biopsy, this approach facilitates simplified, minimally invasive longitudinal monitoring of patients before and during treatment, providing insights into molecular dynamics and treatment response over time.

In this study, we hypothesize that an integrative multi-omics analysis of circulating cfDNA methylation and EVs-derived miRNAs obtained from liquid biopsy can reveal molecular subtypes of NSCLC associated with distinct survival phenotypes. By leveraging the complementary regulatory information provided by epigenetic and miRNA-mediated layers, our approach aims to enhance patient stratification and provide clinically relevant insights into the biological mechanisms associated with differences in overall survival among NSCLC patients receiving immunotherapy.

## Methods

### Patient cohort

This study included a cohort of 79 patients diagnosed with metastatic NSCLC who received ICIs, either as monotherapy (pembrolizumab or atezolizumab) or in combination with chemotherapy. Patients were recruited between February 2015 and May 2023 at two hospitals in Spain: Hospital Universitario Regional de Málaga and Hospital Universitario Virgen de la Victoria. Peripheral blood samples were collected longitudinally at multiple time points: prior to ICI initiation (T1, n = 71), after the second treatment cycle (T2, n = 65), at six months (T3, n = 41), and at twelve months (T4, n = 29). Not all patients had paired samples available (Supplementary Figure S2).

### Inclusion and exclusion criteria

Patients were eligible for inclusion in the study if they met all the following criteria:

I. histologically or cytologically confirmed diagnosis of advanced NSCLC (stage IIIB or IV NSCLC);
II. age ≥18 years at the time of consent;
III. initiation of treatment with ICIs, either as monotherapy (pembrolizumab or atezolizumab) or in combination with platinum-based chemotherapy;
IV. adequate organ function, defined by standard clinical laboratory thresholds for hematologic, renal, hepatic, and cardiac parameters as determined by institutional guidelines.

Exclusion criteria were as follows:

I. presence of concurrent malignancies or a history of other neoplastic diseases within the last 5 years, excluding non-melanoma skin cancer or in situ cervical carcinoma;
II. diagnosis of any autoimmune or inflammatory disorder requiring systemic immunosuppression;
III. active, uncontrolled infections at the time of enrollment;
IV. known contraindications or hypersensitivity to any components of the ICI or chemotherapy regimens;
V. receipt of any prior systemic oncologic treatment, including chemotherapy, targeted therapies, or immunotherapy, for advanced disease; and
VI. inability or unwillingness to provide written informed consent.

All patients were enrolled prospectively based on these criteria. Clinical, pathological, and treatment-related data were extracted from comprehensive electronic health records, ensuring completeness and no loss to follow-up across all study timepoints. Primary resistance was defined according to SITC criteria ^15^ as disease progression occurring within six months of initiating ICI therapy, following the administration of at least two complete treatment cycles. In contrast, non-resistant patients exhibited either a tumor response or stable disease, as determined by RECIST v1.1, sustained for ≥ six months.

### Ethical approval

The study protocol adhered to the Declaration of Helsinki and received approval from the Comité de Ética de la Investigación Provincial de Málaga (Approval date: October 26, 2017; Project Title: “Omics integration for precision cancer immunotherapy,” ID: 799818, H2020-MSCA-IF-2017). Written informed consent was obtained from all participants.

### cfDNA methylation profiling - cfMeth

cfDNA was extracted from 3 mL of frozen plasma per time point using QIAamp Circulating Nucleic Acid kit (Qiagen, 55114) and NEBNext Enzymatic Methyl-seq (EM-seq) protocol (New England Biolabs) was used for profiling DNA methylation at single-base resolution. EM-seq enables detection of both 5-methylcytosine (5mC) and 5-hydroxymethylcytosine (5hmC) ^16^. A custom target panel was developed, comprising 344 genes (including 2,276 promoter regions) and 841 enhancers, based on our previous report on a transcriptomic signature of response and prognosis in ICIs treated patients ^17^. Paired-end sequencing was conducted using an Illumina NovaSeq system (S4 200 bp kit, 10% PhiX spike-in) to achieve a coverage depth of 500X. Sequencing data were subjected to adapter trimming and quality filtering with Trim Galore (Babraham Bioinformatics Institute). Reads were aligned to the human genome (hg38) using Bismark ^18^, allowing discrimination of unmethylated cytosines. Duplicate reads were removed, and methylation/hydroxymethylation calls were generated using‘bismark_methylation_extractor‘. The Picard tool‘CollectHsMetrics‘was used to evaluate hybrid selection metrics based on the custom panel. Hereafter, this omic layer will be designated as cfMeth throughout the study.

### EVs-miRNA profiling - miRNA

Extracellular vesicles were isolated from 1 mL of cryopreserved plasma using the ExoEasy Midi Kit (Qiagen), and total RNA was extracted using the ExoRNeasy Midi Kit (Qiagen), following the manufacturer’s instructions. RNA quality and quantity were assessed using the Agilent 2100 Bioanalyzer. Small RNA libraries were prepared with the NEBNext Multiplex Small RNA Library Prep Kit and sequenced with Illumina Novaseq 6000 platform using single-end reads with a yield of ∼ 10 M reads per sample. The miRNA sequencing data were processed following the ENCODE standards. Quality control, alignment, and annotation were performed using the COMPSRA pipeline ^19^. From this point forward, this omic dataset will be referred to as miRNA.

### Single-Omics analyses

Differential Expression of miRNAs: Differential expression analysis of EV-derived miRNAs was carried out using the DESeq2 package ^20^. miRNAs with an adjusted p-value < 0.05 and |log₂ fold change| > 1.5 were considered significant. Raw counts from COMPSRA were normalized, and expression patterns were visualized via heatmaps and hierarchical clustering. Functional enrichment of differentially expressed miRNAs was performed using miEAA 2.0 for Gene Ontology pathways ^21^.

Differential Methylation and Hydroxymethylation Analysis: Differentially regions analysis were identified using the MethylKit R package ^22^. Analyses considered genomic features including CpG islands, enhancers, shores, shelves, and transcription factor binding sites. Filtering criteria included minimum coverage of 10X and at least 50% of CpG sites per region covered across samples. A multinomial overdispersion model was applied to account for variability in coverage and complex biological effects. Group comparisons were performed using a Chi-squared test, with p-values adjusted using the Benjamini-Hochberg method. Sex and batch were included as covariates. Functional enrichment analysis was performed using the enrichGO function from the clusterProfiler R package ^23^. In the analysis of cfMeth, no distinction was made between 5mC (methylation) and 5hmC (hydroxymethylation); therefore, the observed differences reflect the combined signal of both modifications (5modC).

Both methylation and miRNA datasets were quantile normalized prior to downstream integration.

### Multi-omics integration

Multi-omics integration was conducted using Multi-Omics Factor Analysis (MOFA2) ^24^, a latent factor model that handles missing data across layers (not all patients had complete data across all omics layers). MOFA2 is an unsupervised computational framework that identifies latent factors capturing the principal sources of variability across multiple omics layers. A factor represents a latent variable. The MOFA2 model infers these factors, enabling the decomposition of high-dimensional molecular data into interpretable, lower-dimensional components. These latent factors can subsequently be used to stratify samples into molecular subtypes, each defined by distinct multi-omics signatures. Two independent MOFA2 models were constructed for time points T1 and T2. Prior to integration, feature selection was performed based on univariate Cox proportional hazards models for OS and progression-free survival (PFS). Only molecular features (cfMeth and miRNAs) with p < 0.05 in either OS or PFS models were retained. Model hyperparameters, including the number of latent factors (range: 2–20), were tuned to optimize survival stratification. Clustering was conducted using Partitioning Around Medoids (PAM) for 2 to 4 clusters. The best-fitting model was selected based on the log-rank p-value and median OS and PFS among identified groups. The clusters generated from the MOFA2 models were referred to as MOFA-derived clusters (MDC). Each time point generated its own molecular classification: MDC-T1 and MDC-T2, representing the MOFA-derived molecular signatures present at baseline and after the second treatment cycle, respectively. To assess the temporal stability of the learned molecular signatures, the latent factors inferred from the T1 and T2 MOFA2 models were projected onto the remaining samples from T1 to T4.

cfMeth and miRNAs, due to their high dimensionality and complex structure, were prioritized for integration. In contrast, flow cytometry data—considered low-dimensional—were excluded from the integrative modeling but were used in downstream analyses to explore biological correlations with the MOFA-derived factors and clusters.

### Flow cytometry analysis

Peripheral blood mononuclear cells (PBMCs) were analyzed from 10 mL of whole blood at all time points using a BD FACS Canto II flow cytometer (BD Biosciences, San Jose, CA, USA). Four antibody panels were used to characterize immune cell subsets. The following fluorochrome-conjugated anti-human antibodies were used for immunophenotyping:

- Panel 1: PE/Cyanine7 anti-CD3 (BioLegend, Cat# 344816), FITC anti-CD8a (BioLegend, Cat# 301006), APC anti-PD-1 (CD279) (BioLegend, Cat# 621610), and PE anti-PD-L1 (CD274, B7-H1) (BioLegend, Cat# 393608).
- Panel 2: FITC anti-CD4 (BioLegend, Cat# 317408), APC anti-CD25 (BioLegend, Cat# 302610), and PE anti-CD127 (IL-7Rα) (BioLegend, Cat# 351304).
- Panel 3: PE anti-CD56 (NCAM) (BioLegend, Cat# 318306) and PE/Cyanine7 anti-CD16 (BioLegend, Cat# 360708).
- Panel 4: APC anti-CD19 (BioLegend, Cat# 302212).

Data were acquired using DIVA software v9.0 and further analyzed with the flowTOTAL pipeline^25^ to obtain the following gating strategy:

- Panel 1: CD3⁺ T cells (CD3+/Lymphocytes), cytotoxic CD8⁺ T cells (CD8+/CD3+), CD3⁺PD1⁺ T cells (CD279+/CD3+), CD3⁺PD-L1⁺ T cells (CD274+/CD3+), CD8⁺PD1⁺ T cells (CD279+/CD3+CD8+) and CD8⁺PD-L1⁺ T cells (CD274+/CD3+CD8+).
- Panel 2: CD4⁺ T cells (CD4+/Lymphocytes) and regulatory T cells-Tregs (CD25+CD127-/CD4+).
- Panel 3: Natural killer cells (CD56+CD16-/Lymphocytes) and cytotoxic NK cells (CD56+CD16-/Lymphocytes).
- Panel 4: B cells (CD19+/Lymphocytes).

### External validation

To explore the generalizability of the identified molecular signatures, external validation was pursued despite the absence of public datasets that simultaneously profile both cfDNA methylation and EV-derived miRNAs in NSCLC patients treated with ICIs. Acknowledging this limitation, validation was conducted independently for each omic layer using publicly available, clinically relevant datasets.

For miRNA validation, we used the dataset published by Genova et al., 2024 ^26^ (GSE207715), which includes EV-derived miRNA expression profiles from baseline liquid biopsies of 54 NSCLC patients prior to initiation of ICI therapy. The miRNA-based molecular signatures identified in our study at both T1 and T2 were validated in this external cohort. Both PFS and OS information were available. For cfDNA methylation signatures, external validation was performed using the SMC Cohort ^27^ (GSE119144), which includes DNA methylation profiles from tumor tissue biopsies of 57 NSCLC patients obtained prior to initiation of ICI therapy. Methylation data were generated using the Illumina Infinium HumanMethylation850 BeadChip array. In this case, only PFS information was available.

Technical limitations of both external datasets should be considered when interpreting the validation results. The Genova et al., 2024 cohort was generated using an array-based technology, and therefore the miRNA species profiled do not completely overlap with those captured by the next-generation small RNA sequencing used in our study. For cfMeth validation, the SMC Cohort comprises tumor tissue samples rather than plasma-derived cfDNA, introducing a biological distinction in sample origin. Moreover, while our data were obtained through targeted profiling of all CpG sites of candidate genes and genomic regulatory regions such as enhancers, the SMC Cohort assesses methylation at representative individual CpG sites across the HumanMethylation850 space.

To overcome these platform disparities, we systematically identified and extracted only the features that overlapped between our datasets and each respective validation cohort.

### Machine Learning-Based MDC for external validation

To enable external validation of MDC labels in independent datasets it was necessary to construct predictive models capable of assigning MDC-T1 and MDC-T2 labels based solely on miRNA expression data and in parallel with cfDNA methylation levels. Four independent supervised classifiers were developed using the training data. For each time point (T1 and T2) and omics (cfMeth and miRNA), Random Forest (RF), radial support vector machines (SVM-Radial) and Lasso logistic regression models were trained using stratified 5-fold cross-validation, with the goal of maximizing predictive performance; AUC (area under the ROC curve). After evaluating performance across folds, the model with the highest AUC was selected. The best-performing model was then applied to the Genova cohort and SMC Cohort, independently, to assign MDC labels (predicted-MDC-T1 and predicted-MDC-T2). Survival analyses in this cohort were then stratified according to the predicted MDC groups.

### Statistical analyses

All statistical analyses were performed using R (v4.2.0; RRID:SCR_001905). Continuous variables were summarized as medians and compared using Wilcoxon rank-sum tests; categorical variables were described as counts and percentages. For comparisons between categorical variables, Chi-squared or Fisher’s exact tests were applied as appropriate based on event counts. Kaplan–Meier survival curves were generated for OS and PFS, with group comparisons made using the log-rank test. A schematic of the overall methodological workflow is provided in Supplementary Figure S1.

## Results

### Clinical characterization and multi-omic model construction

A total of 79 patients with advanced NSCLC were enrolled in this study (Table 1). Among them, 55 had adenocarcinoma and 24 had non-adenocarcinoma tumors. Patients were treated with ICIs, either as monotherapy (n=26) or in combination with platinum-based chemotherapy (n=53). The cohort included 60 males and 19 females, with a mean age of 65 years (SD=7.5). Based on tumor PD-L1 expression, 59 patients had Tumor Proportion Score (TPS) < 50%, while 16 exhibited PD-L1 levels ≥ 50%. Clinical evaluation identified 32 patients as having primary resistance to ICI therapy—defined according to SITC guidelines ^15^—while the remaining 47 were considered non-resistant.

**Table 1.** Clinicopathological characteristics of the NSCLC patient cohort included in the study (n=79).

| Variables | Categories | Statistic | Missing values |
| --- | --- | --- | --- |
| Age |  | 65.4 (48.34-84.9 ) | 0 (0%) |
| Sex |  |  | 0 (0%) |
|  | *Female* | 19 (24.05%) |  |
|  | *Male* | 60 (75.95%) |  |
| Baseline ECOG Performance Status |  |  | 3 (3.8%) |
|  | 0 | 18 (22.79%) |  |
|  | 1 | 56 (70.89%) |  |
|  | 2 | 2 (2.54%) |  |
| Baseline LDH | Normal | 57 (72.15%) | 15 (18.99%) |
|  | High | 7 (8.86%) |  |
| PD-L1 Expression Group | <50% TPS | 59 (74.68%) | 4 (5.06%) |
|  | >50% TPS | 16 (20.26%) |  |
| Pulmonary Metastasis | No | 46 (58.23%) | 1 (1.27%) |
|  | Yes | 32 (40.50%) |  |
| Bone Metastasis | No | 57 (72.15%) | 1 (1.27%) |
|  | Yes | 21 (26.58%) |  |
| Hepatic Metastasis | No | 70 (88.60%) | 1 (1.27%) |
|  | Yes | 8 (10.13%) |  |
| Lymph Node Metastasis | No | 36 (45.57%) | 1 (1.27%) |
|  | Yes | 42 (53.16%) |  |
| Cutaneous Metastasis | No | 49 (62.03%) | 1 (1.27%) |
|  | Yes | 29 (36.70%) |  |
| Brain Metastasis | No | 70 (88.60%) | 1 (1.27%) |
|  | Yes | 8 (10.13%) |  |
| Diagnosis Subtype | Adenocarcinoma | 55 (69.62%) | 0 (0%) |
|  | Non-Adenocarcinoma | 24 (30.38%) |  |
| Type of Immunotherapy Treatment | IT | 26 (32.91%) | 0 (0%) |
|  | QT+IT | 12 (15.19%) |  |
|  | QT+IT+Others | 41 (51.90%) |  |
| Primary Resistance to Immunotherapy | Resistant | 32 (40.51%) | 0 (0%) |
|  | Non-resistant | 47 (59.49%) |  |
| Maximum Toxicity Grade |  | 1.14 (0-5 ) | 7 (8.86%) |
| Immunotherapy-Related Toxicity | No | 34 (43.04%) | 1 (1.27%) |
|  | Yes | 44 (55.7%) |  |
| OS Status | 0 | 17 (21.52%) | 0 (0%) |
|  | 1 | 62 (78.48%) |  |
| PFS Status | 0 | 18 (22.78%) | 0 (0%) |
|  | 1 | 61 (77.22%) |  |
| OS |  | 15.41 (CI:10.23-24.36) |  |
| PFS |  | 10.92 (CI:7.87-17.64) |  |

\*\*\* Continuous variables are presented as median values with ranges. Qualitative data are shown as both absolute and relative frequencies. Survival data for overall survival (OS) and progression-free survival (PFS) are reported as median survival times with corresponding confidence intervals.

At baseline (T1), 71 plasma samples were obtained. Of these, 50 patients had matched cfDNA methylation (cfMeth) and EVs-derived miRNA (miRNA) data, while 2 had only cfMeth and 19 had only miRNA data. At the post-cycle time point (T2), 65 samples were available, with 47 containing both omics layers, 4 with cfMeth only, and 14 with miRNA only (Supplementary Figure S2-A). Both cfMeth and miRNA datasets, considered high-dimensional molecular data types, were included in the multi-omic integration framework.

Univariate Cox regression identified molecular features associated with OS or PFS to be included in MOFA2 modeling. At T1, 136 miRNAs and 298 cfMeth features met the significance threshold (p < 0.05) (Supplementary Table S1), while at T2, 81 miRNAs and 714 cfMeth features were retained (Supplementary Table S2). These filtered variables served as the input for the construction of independent MOFA2 models for T1 and T2. The number of latent factors for each model was selected based on optimization of model fit, evaluated through total variance explained and the ability of downstream clusters to stratify patients by overall and progression-free survival. The number of selected factors were 3 and 5 respectively (Supplementary Figure S3).

In the T1 model (Figure 1-A), MOFA2 explained 23% of the variance in miRNA data and 25% in cfMeth data. Analysis of latent factors revealed that Factor 1 (F1) captured contributions from both omic layers, with a stronger weight from cfMeth (∼15%) compared to miRNAs (∼10%). Factor 2 (F2) showed a shift toward a greater influence from miRNAs, while Factor 3 (F3) demonstrated balanced contributions between both modalities. In contrast, the T2 model (Figure 1-B) revealed more pronounced differences in total variance explained across layers, with cfMeth accounting for 50% and miRNAs for 25%. F1 displayed a similar distribution of contributions as in T1, whereas F2 was predominantly driven by miRNAs and F3 by cfMeth. Factors 4 (F4) and 5 (F5) exhibited more symmetric input from both data types.

**Figure 1.**
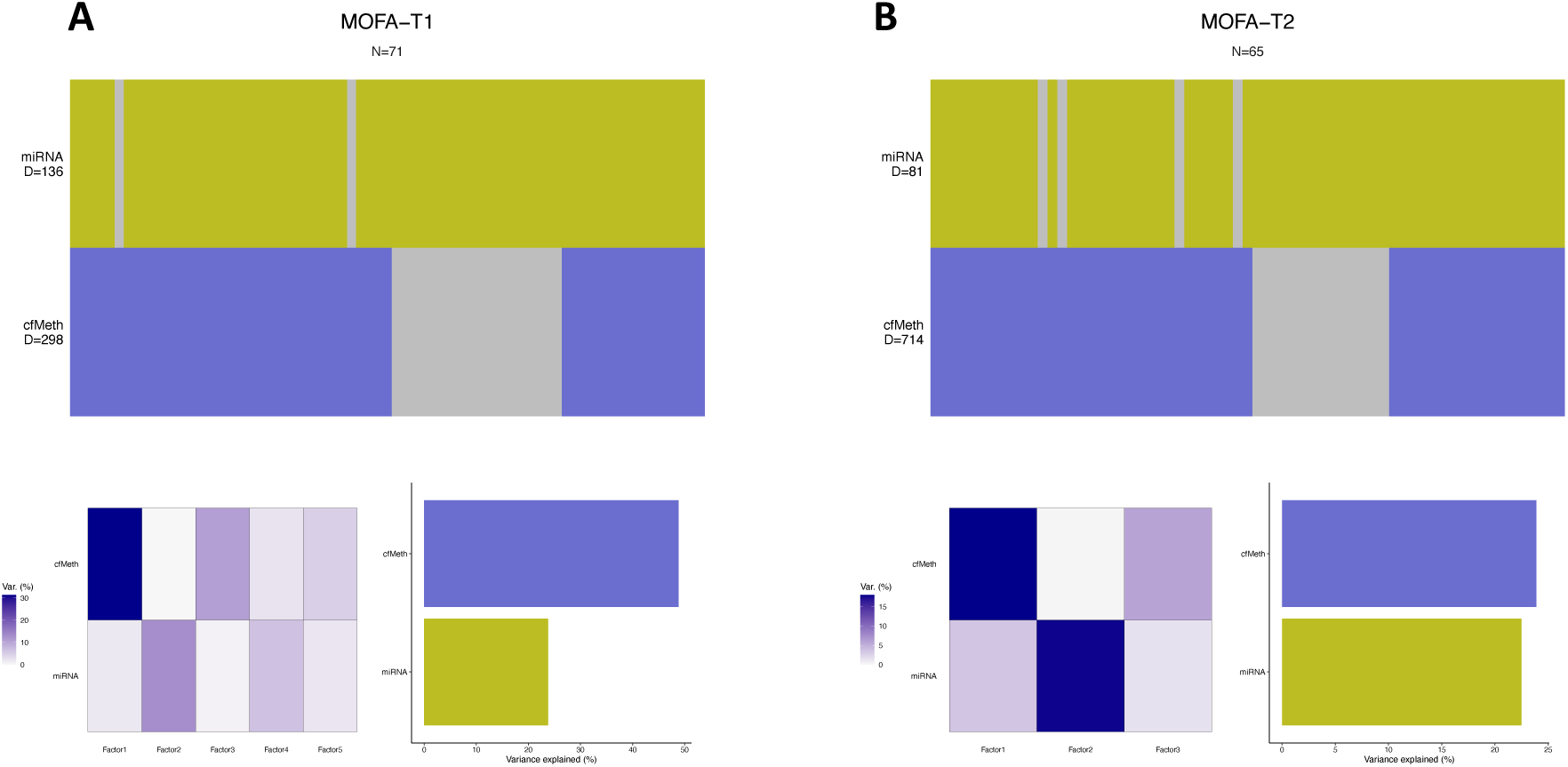
Overview of multi-omic integration and latent factor composition for T1 and T2 models. (A) T1 model: Schematic representation of the sample composition used for MOFA2 integration (n=71). Samples are color-coded to indicate paired miRNA and cfMeth data or presence of missing omic layers (in grey). The barplot shows the proportion of total variance explained by each omic layer, and the heatmap the variance captured by each latent factor. (B) T2 model: Equivalent schematic for T2 (n=65), displaying sample pairing status and variance explained by omic and factor.

These results indicate that the relative importance of each omic layer shifts over time, suggesting dynamic remodeling of the tumor and immune environment under ICI treatment, examined during the individual characterization of the omic layer. The latent factors extracted through MOFA2 serve as the foundation for downstream clustering and survival stratification analyses.

### MOFA2 identifies three latent molecular subtypes with distinct clinical outcomes

To stratify patients based on integrative molecular profiles, unsupervised clustering using the PAM algorithm was applied to the latent factors derived from the MOFA2 models at both T1 and T2. Optimization of cluster number based on internal clustering metrics and survival separation consistently revealed three robust molecular subgroups per time point. These groups were designated as MDC1-T1, MDC2-T1, and MDC3-T1 for the baseline model (T1), and MDC1-T2, MDC2-T2, and MDC3-T2 for the post-treatment model (T2).

A heatmap illustrating the expression and methylation levels of the features selected for integration, alongside the MDC annotations, is shown in Figure 2-A. In the T1 model, MDC1-T1 was characterized by elevated scores in F1 and F3, while MDC3-T1 exhibited a strong inverse association with F2. MDC2-T1 presented an intermediate profile, showing similarities to MDC3-T1 in F1 and F3, and to MDC1-T1 in F2 (Figure 2-B). Variable loadings per factor are depicted in Figure 2-C, showing that F2 was primarily driven by miRNAs, with minor contribution from cfMeth, whereas F1 and F3 exhibited stronger integration of both omic layers.

**Figure 2.**
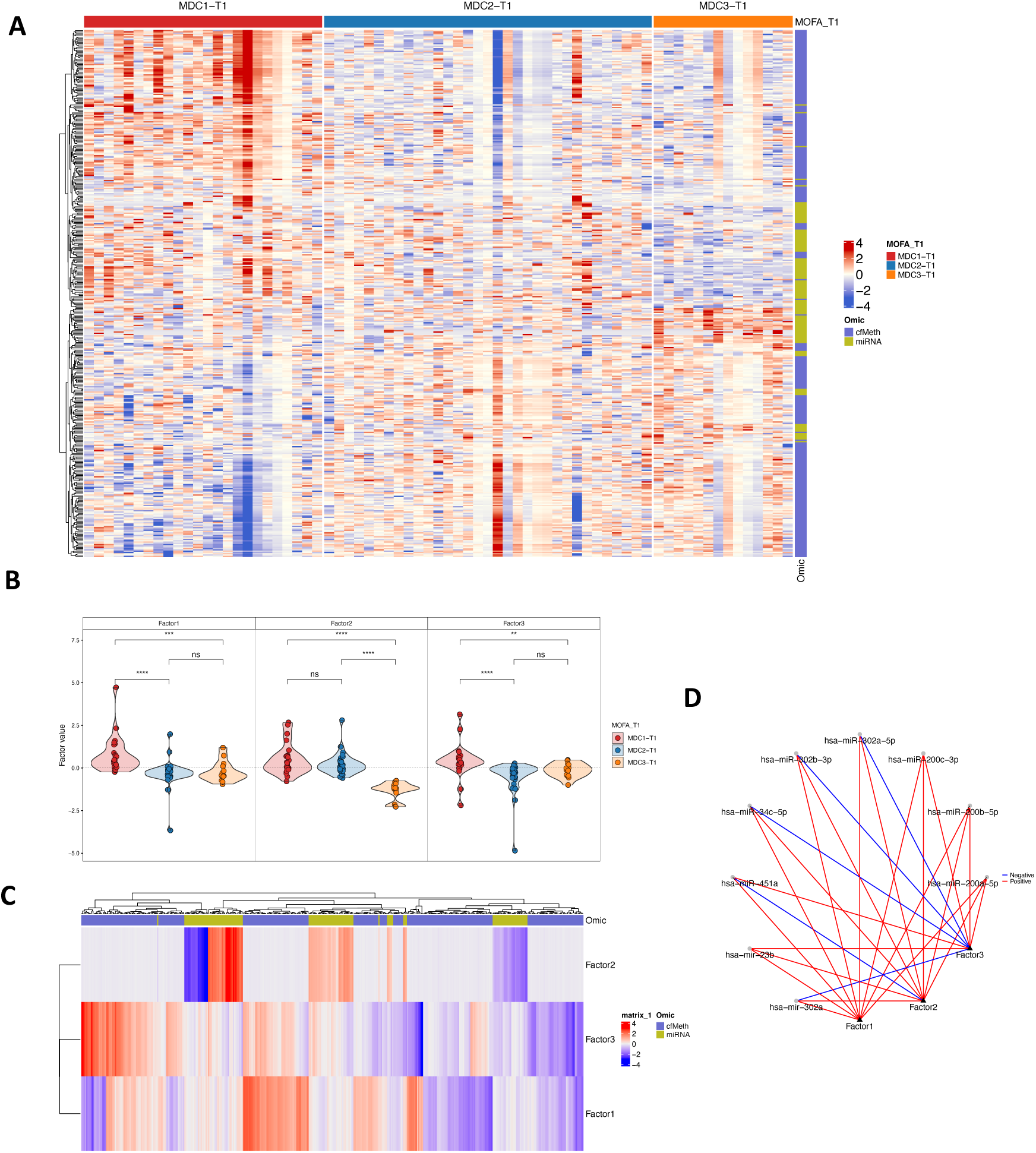
Integrative latent structure and multi-omic feature architecture of the T1 MOFA2 model. (A) Heatmap of scaled molecular features (miRNAs and cfMeth regions) used in the MOFA2 model at T1. Columns represent patient samples and rows denote selected features. Color annotations at the top indicate MDC-T1 cluster assignments (MDC1-T1, MDC2-T1, MDC3-T1). (B) Violin plots showing the distribution of latent factor values across the three MDC-T1 subtypes. Statistical comparisons were performed using Wilcoxon tests; significance levels are indicated as follows: ***p < 0.001; **p < 0.01; *p < 0.05; ns: not significant. (C) Heatmap of feature loadings across latent factors. Rows represent MOFA2 factors (F1-F3) and columns represent molecular features. Color scale indicates loading weight, highlighting omic-specific contributions. (D) Network representation of molecular features and their associated latent factors. Edges connect features to factors when loading weights above the defined threshold in all three factors.

To further explore the interaction between molecular features and latent factors, weight matrices were transformed into a network, filtering for features contributing to all three factors (Figure 2-D). This network revealed a core module of 9 EV-derived miRNAs. Among them, *hsa-miR-23b, hsa-miR-200c-3p, hsa-miR-200b-5p*, and *hsa-miR-200a* showed positive loadings across all factors. In contrast, *hsa-miR-302a, hsa-miR-34c-5p, hsa-miR-302-3p,* and *hsa-miR-302-5p* were positively associated with F1 and F2 but negatively with F3, whereas *hsa-miR-451a* exhibited the opposite pattern.

In the T2 model, cluster structure was again resolved into three distinct MDCs (Figure 3-A). MDC1-T2 displayed low values in F2 and intermediate levels in F4, while MDC2-T2 was defined by high activity in F4. MDC3-T2 showed reduced contribution from both F1 and F4 (Figure 3-B). The variable loading map in Figure 3-C demonstrated, as in T1, limited cfMeth involvement in F2, with greater omic integration in the remaining latent components. Network analysis under strict criteria produced no nodes meeting all-factor thresholds. When relaxed to features contributing to at least three factors, a reduced network emerged with two interconnected communities: one comprising F1, F3, F4, and F5, and another centered on F2. These were bridged by four key miRNAs*: hsa-miR-6873-3p, hsa-miR-539-5p, hsa-miR-615-3p,* and *hsa-miR-1185-2-3p* (Figure 3-D).

**Figure 3.**
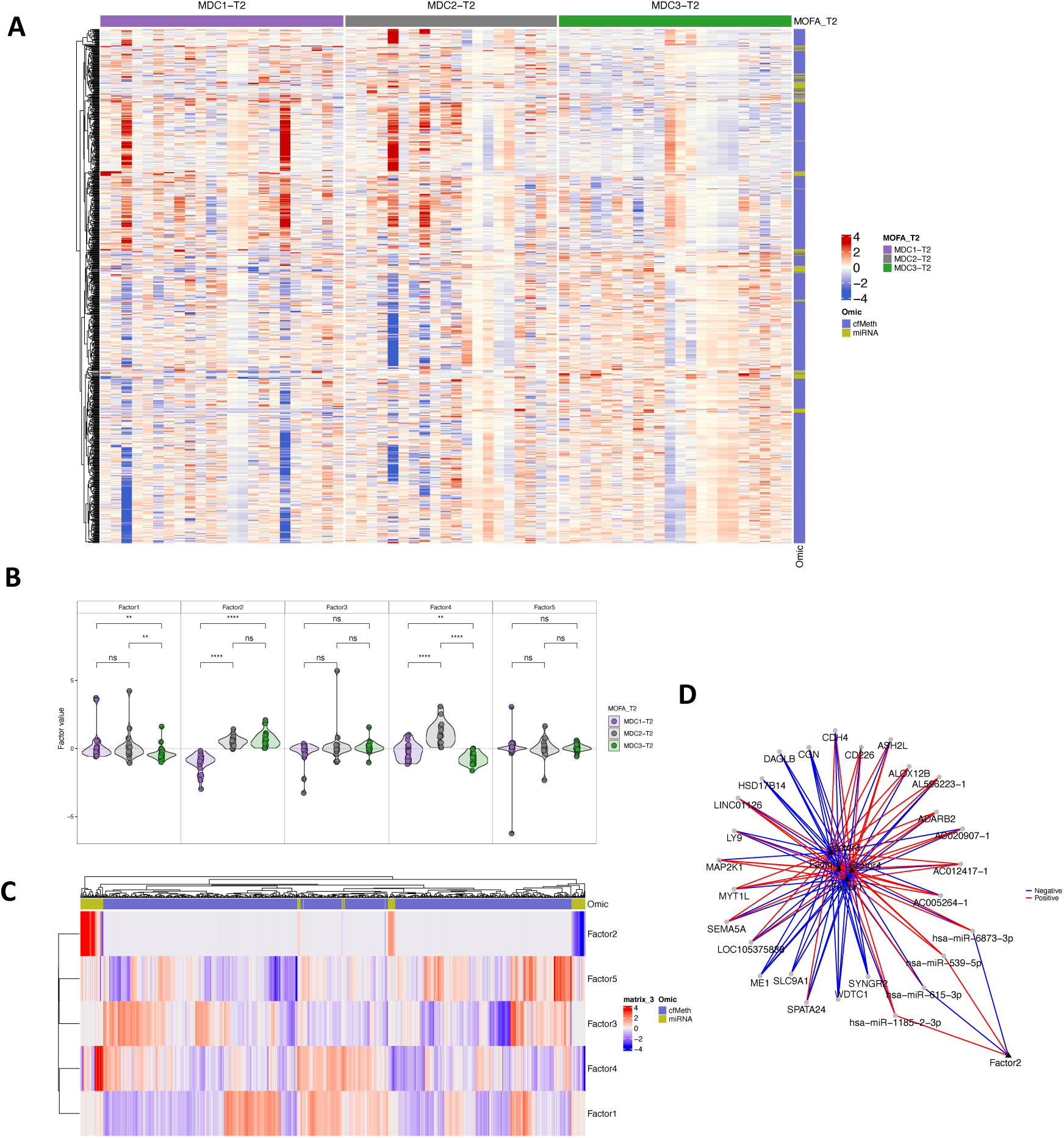
Integrative latent structure and multi-omic feature architecture of the T2 MOFA2 model. (A) Heatmap of scaled molecular features integrated in the MOFA2 model at T2. Columns represent patient samples and rows correspond to selected variables. Top annotation indicates MDC-T2 subtype assignments (MDC1-T2, MDC2-T2, MDC3-T2). (B) Violin plots comparing the distribution of latent factor values across MDC-T2 subtypes. Wilcoxon tests were used for pairwise comparisons. Significance is denoted as ***p < 0.001; **p < 0.01; *p < 0.05; ns: not significant. (C) Heatmap of variable loadings across the five MOFA2 latent factors. Rows represent factors (F1-F5), and columns denote molecular features. Color gradient reflects the contribution strength of each feature to each factor. (D) Network representation of molecular features and their association with latent factors. Edges indicate features with loading weights above the defined threshold in at least three factors.

To further interpret the biological drivers of each MDCs, we identified the top-weighted molecular features per factor in both models (Supplementary Table S3). In the T1 model, F1 was primarily driven by *hsa-miR-302a-5p, hsa-miR-200c-3p* and two cfDNA methylation regions located at chr7:2,567,162-2,568,844 (*IQCE*) and chr19:42,202,165-42,204,413 (*DEDD2*), all with positive loadings. F2 was characterized by *hsa-miR-199a-3p* and a methylation region at chr3:52,409,785-52,409,793 (*BAP1*), while F3 showed inverse contribution from *hsa-miR-302a-5p* and methylation regions at chr17:7,445,553-7,445,983 (*FGF11*) and chr17:48,611,037-48,611,105 (*HOXB7*), while positive loading in *hsa-miR-200c-3p*. In the T2 model, F1 was dominated by *hsa-miR-1185-2-3p* and a cfMeth region at chr2:43,224,342-43,225,700 (*ZFP36L2*). F2 showed strong contributions from *hsa-miR-106b-5p*, with associated methylation regions at chr7:159,030,625-159,030,782 (*VIPR2*); positively, and chr17:78,130,335-78,130,622 (*TMC6, TMC8*); negatively. F3 included *hsa-miR-1273c* and a locus at chr8:98,948,226-98,948,318 (*OSR2*), while F4 was negatively loaded by *hsa-miR-30a-5p* and a region at chr4:152,322,939-152,323,048 (*FBXW7*). Finally, F5 was inversely driven by *hsa-miR-1185-2-3p* and positively associated with methylation at chr21:38,400,473-38,400,597 (*ERG*).

Survival analysis revealed significant differences in clinical outcomes across MDCs (Figure 4). At T1, MDC1-T1 was associated with the poorest prognosis, showing median PFS and OS of 4.54 and 7.64 months, respectively. MDC2-T1 had intermediate outcomes (median PFS and OS of 12.23 and 19.71 months), whereas MDC3-T1 demonstrated the most favorable survival, with median PFS and OS of 28.89 and 40.79 months. In the T2 model, while three clusters were identified, the survival landscape differentiated into two distinct risk groups: MDC1-T2 and MDC2-T2 formed a low-survival state; 4.66 and 7.71 months in median PFS whereas 7.78 and 9.9 months respectively in OS; while MDC3-T2 represented a high-survival group (28.89 and 40.79 months median survival, PFS and OS).

**Figure 4.**
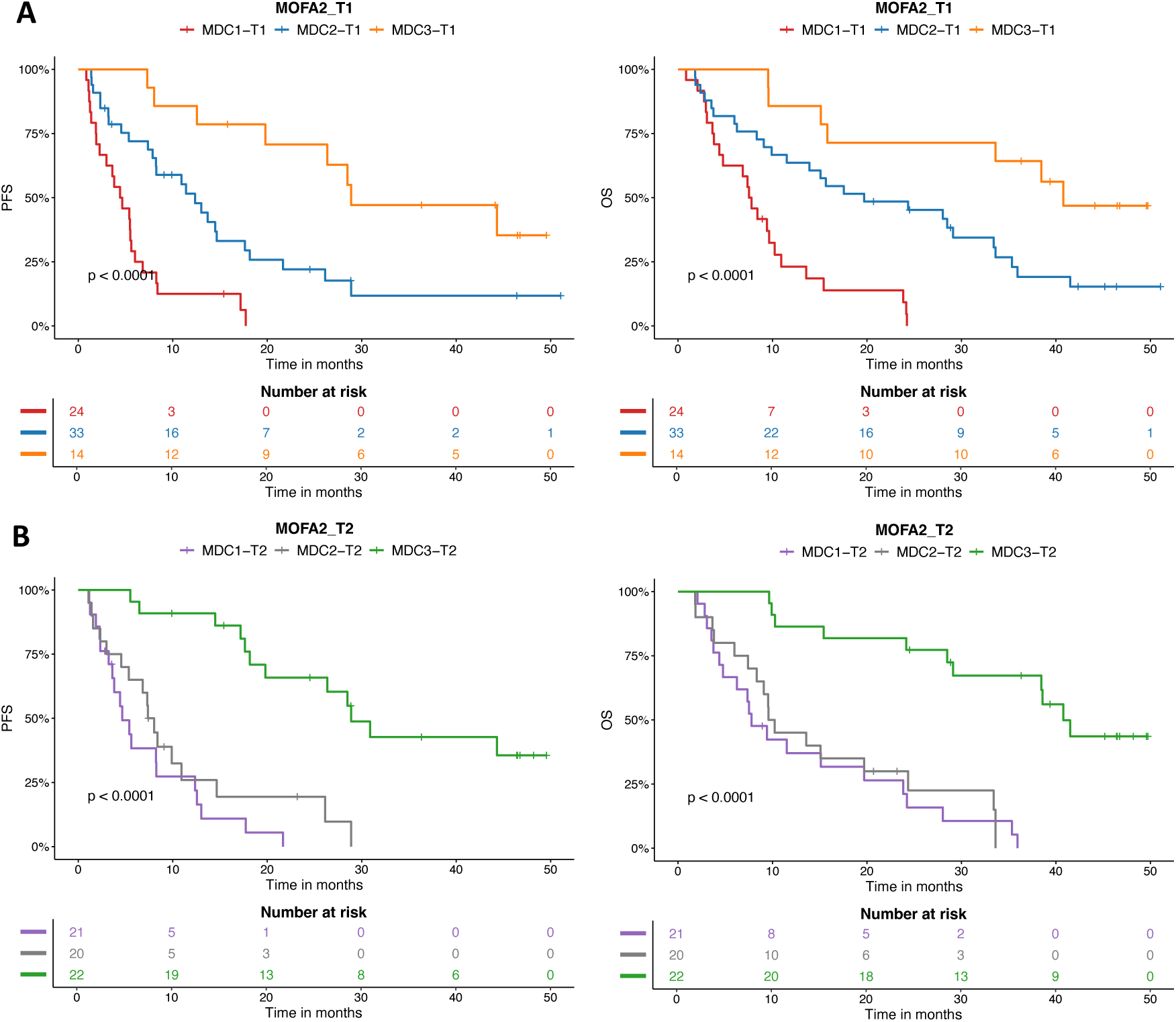
Survival analysis by MOFA2-derived molecular subtypes (MDCs) at baseline (T1) and early treatment (T2). Kaplan–Meier survival curves showing overall survival (OS) and progression-free survival (PFS) for patients stratified by MDC clusters derived from MOFA2 integration. (A) OS and PFS in the baseline model (T1), comparing MDC1-T1, MDC2-T1, and MDC3-T1. (B) OS and PFS in the early on-treatment model (T2), comparing MDC1-T2, MDC2-T2, and MDC3-T2. Statistical significance was assessed using the log-rank test.

These results indicate that integrative molecular profiles at both baseline and early treatment stages uncover latent structures capable of stratifying patients into clinically distinct prognostic subgroups. To investigate the temporal robustness and translational relevance of these molecular programs, we next projected the MOFA2-derived signatures onto independent samples collected at different treatment time points not included in model construction.

### Projection of MOFA-derived clusters onto T1, T2, T3 and T4 samples

To investigate the temporal consistency and clinical relevance of the molecular subtypes identified by MOFA2, we projected the latent space signatures from each integrative model onto samples collected at treatment stages not used in model development. Specifically, MDC-T1 were projected onto samples from T2, T3, and T4, and MDC-T2 onto T1, T3, and T4. This approach leverages the MOFA2 framework, which allows latent factor projection using the learned loading matrices and matrix inversion to perform deconvolution on new data.

In the T1 model, dimensionality reduction of the projected latent space using UMAP revealed that samples from different time points integrated cohesively into the projected space without forming isolated clusters, suggesting continuity in the model structure across time. Clusters MDC2-T1 and MDC3-T1 retained distinct grouping patterns, whereas MDC1-T1 appeared more heterogeneous, splitting into two partially overlapping regions (Figure 5-A). Survival analyses based on these projected cluster labels at later time points did not yield statistically significant differences in OS (Figure 5-B) or PFS (Supplementary Figure S4-A), although a weak association was observed for OS at T3 (p ≈ 0.051), where MDC1-T1 consistently exhibited the poorest prognosis.

**Figure 5.**
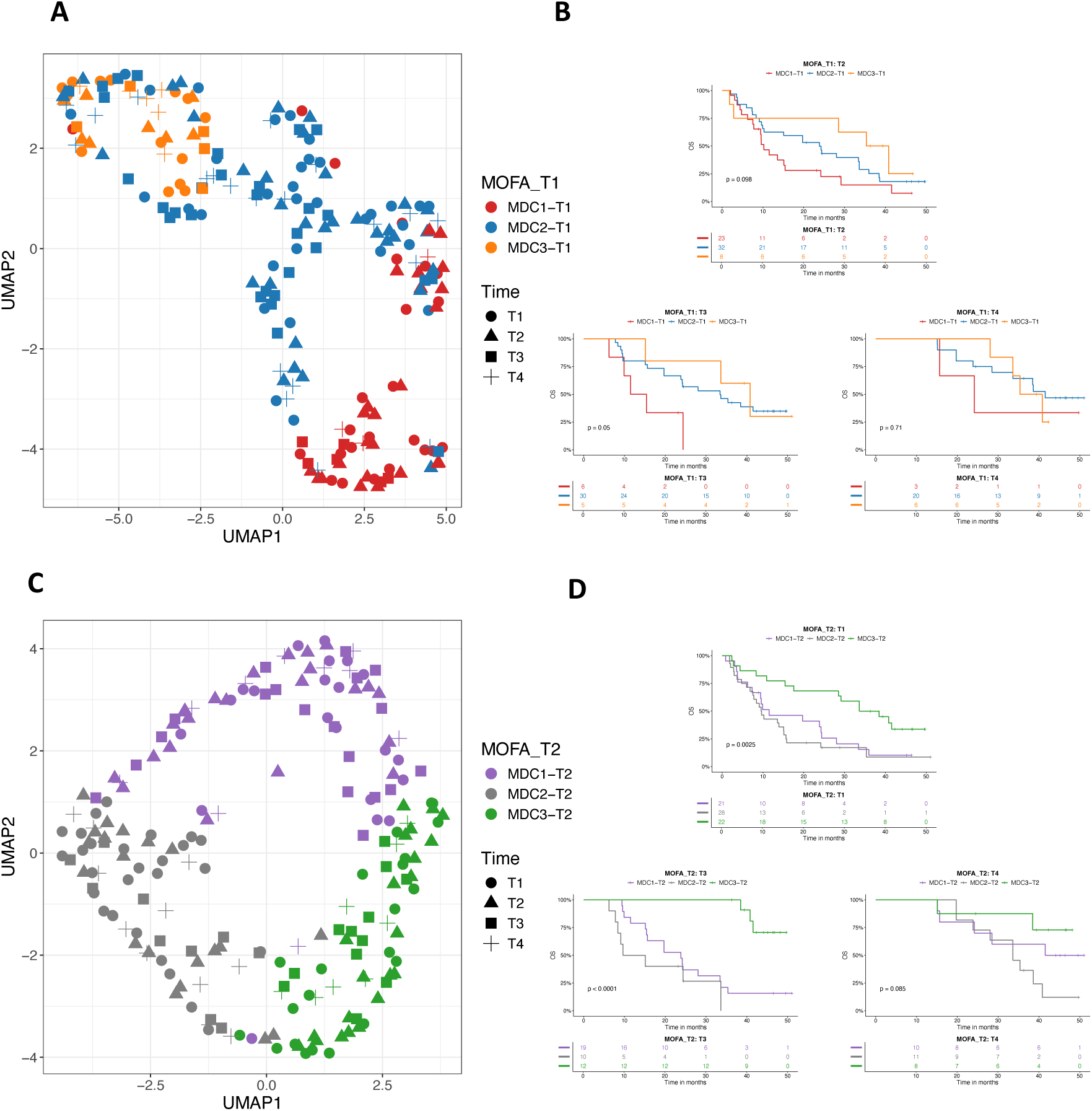
Longitudinal projection of MDCs stratifies NSCLC patients in OS. (A) UMAP embedding of projected samples from T1, T2, T3, and T4 using the MDC-T1 model. Samples are colored according to MDC-T1 subtype assignments and shaped by collection time point. (B) Kaplan– Meier curves for OS of MDC-T1 subtypes projected onto T2, T3, and T4 samples. (C) UMAP of projected T1, T3, and T4 samples based on the MDC-T2 model. (D) Kaplan–Meier curves for OS in projected MDC-T2 subtypes in T1, T3, and T4 samples. Survival differences were evaluated using the log-rank test.

In contrast, the T2 model demonstrated more robust projection behavior. When latent signatures from MDC-T2 were projected onto samples from T1, T3, and T4, UMAP analysis showed well-separated clusters corresponding to MDC-T2 labels, while no clear segregation by time point was observed—indicating that the molecular structure captured by T2 signatures may reflect treatment-adaptive states rather than temporal stage (Figure 5-B). Survival analysis revealed significant separation, log-rank p < 0.05: MDC-T2 projected onto T1 distinguished patients by both OS (Figure 5-D) and PFS (Supplementary Figure S4-B), and T3 projection maintained significance for OS and approached significance for PFS (p = 0.06). In all projections, MDC1-T2 and MDC2-T2 consistently marked poor-outcome groups, while MDC3-T2 was associated with favorable prognosis. No significant survival differences were observed in the T4 projection, however, T4 samples are collected one year after treatment initiation, and survival censoring at this stage may obscure true long-term stratification.

These findings indicate that while baseline-derived molecular subtypes show limited temporal stability, the T2-derived signatures demonstrate greater prognostic consistency across multiple treatment stages, including both pre-treatment and on-treatment samples.

### Molecular characterization of MDC-T1 and MDC-T2 subtypes across omics layers

To define the molecular architecture underlying the MDCs, we performed comprehensive pairwise differential analyses of EV-derived miRNA expression and cfDNA methylation profiles at each time point. Comparisons were conducted across all combinations of MDC subtypes at T1 and T2 to capture both unique and shared biological signatures that may distinguish these latent molecular states (Supplementary Table S4).

In the T1 model (Supplementary Figure S5-A), the comparison between MDC1-T1 and MDC2-T1 revealed a distinct miRNA expression profile, with 22 miRNAs significantly upregulated and two downregulated in MDC1-T1. Expression differences were further amplified in the contrast between MDC1-T1 and MDC3-T1, identifying 32 upregulated and 19 downregulated miRNAs in MDC1-T1. The contrast between MDC2-T1 and MDC3-T1 yielded the most substantial miRNA shift, with 40 miRNAs overexpressed in MDC2-T1 and 32 in MDC3-T1. Notably, intersection of the comparisons involving MDC1-T1 uncovered a conserved six-miRNA signature, with five miRNAs (*hsa-miR-1246, hsa-miR-200c-3p, hsa-miR-429, hsa-miR-200b,* and *hsa-miR-615-3p*) consistently upregulated, and one (*hsa-miR-212-5p*) downregulated. In MDC2-T1, four miRNAs were recurrent across comparisons, including *hsa-miR-133a-3p* (upregulated) and *hsa-miR-1224-5p* and *hsa-miR-1255b-5p* (downregulated), although *hsa-miR-429* showed a reversed direction of change, suggesting context-specific regulation. Among all subtypes, MDC3-T1 demonstrated the most robust and internally consistent miRNA profile, with 37 overlapping differentially expressed miRNAs (22 upregulated, 15 downregulated), reinforcing the biological distinctiveness of this cluster.

At the epigenetic level, the cfDNA methylation analysis further supported the molecular divergence among MDCs. The MDC1-T1 vs. MDC2-T1 comparison identified 31 regions hypermethylated in MDC1-T1 and 280 in MDC2-T1. The comparison between MDC1-T1 and MDC3-T1 revealed six differentially methylated regions, all highly methylated in MDC3-T1 in comparison with MDC1-T1. Notably, both contrasts involving MDC1-T1 shared hypomethylation at loci corresponding to *BLK* and *ADARB2*, suggesting potential subtype-specific regulatory elements. No significant differences were observed between MDC2-T1 and MDC3-T1, indicating that these subtypes may be more transcriptionally than epigenetically divergent at this pre-treatment stage.

In the T2 model (Supplementary Figure S5-B), molecular subtypes continued to exhibit distinct transcriptomic profiles. The comparison between MDC1-T2 and MDC2-T2 identified 35 miRNAs upregulated in MDC1-T2 and 44 downregulated. The contrast between MDC1-T2 and MDC3-T2 yielded 24 miRNAs elevated in MDC1-T2 and 17 suppressed. The molecular distinction between MDC2-T2 and MDC3-T2 was marked by 45 miRNAs enriched in MDC2-T2 and 20 in MDC3-T2. A 20-miRNA core signature, composed of 12 upregulated and 8 downregulated miRNAs, was conserved across comparisons involving MDC1-T2, underscoring the internal coherence of this subtype. Similarly, 28 miRNAs overlapped in MDC2-T2-related comparisons, while MDC3-T2 displayed 11 consistent deregulated miRNAs, most with consistent directional change.

The methylation patterns at T2 revealed fewer, but still informative, differentially methylated regions. The MDC1-T2 and MDC3-T2 comparison yielded 12 regions hypermethylated in MDC1-T2 and 6 in MDC3-T2. MDC2-T2 and MDC3-T2 contrast identified three regions hypermethylated in MDC2-T2 and two in MDC3-T2. No differentially methylated regions were detected between MDC1-T2 and MDC2-T2, suggesting a degree of epigenetic convergence or reduced signal contrast between these subtypes. Furthermore, no recurrent differentially methylated regions were shared across T2 comparisons, indicating that cfDNA methylation profiles at this treatment stage may reflect more compartmentalized and context-specific epigenetic states.

Among the molecular features contributing most consistently to both the single-omic differential analyses and the integrative MOFA2 models, a subset of five miRNAs emerged as particularly informative (Figure 6-A). *hsa-miR-130b-5p* was consistently downregulated in the favorable-outcome cluster MDC3 at both T1 and T2, suggesting the potential bad prognosis tumor-promoting role. Similarly, *hsa-let-7i-5p* showed decreased expression in MDC3 (T1 and T2) and MDC2-T2. In contrast, *hsa-miR-451a* and *hsa-miR-4732-5p* stood out as highly upregulated biomarkers—both were upregulated in MDC3 at T1 and T2 and featured prominently in the latent factor structures of the corresponding MOFA2 models. Finally, *hsa-miR-615-3p* exhibited elevated expression in MDC1-T1 and MDC1-T2, which are associated with poor clinical outcome. At the epigenetic level, two regulatory regions demonstrated consistent subtype-specific methylation patterns linked to survival (Figure 6-B). The chr8:11,550,181–11,550,226 locus, mapped to *BLK*, and the chr10:1,505,491–1,505,535 region, associated with *ADARB2*, were more heavily methylated in subtypes with superior outcomes (MDC3-T1, MDC3-T2, and MDC2-T1), and hypomethylated in subtypes with poorer prognosis (MDC1-T1, MDC2-T2, MDC1-T2).

**Figure 6.**
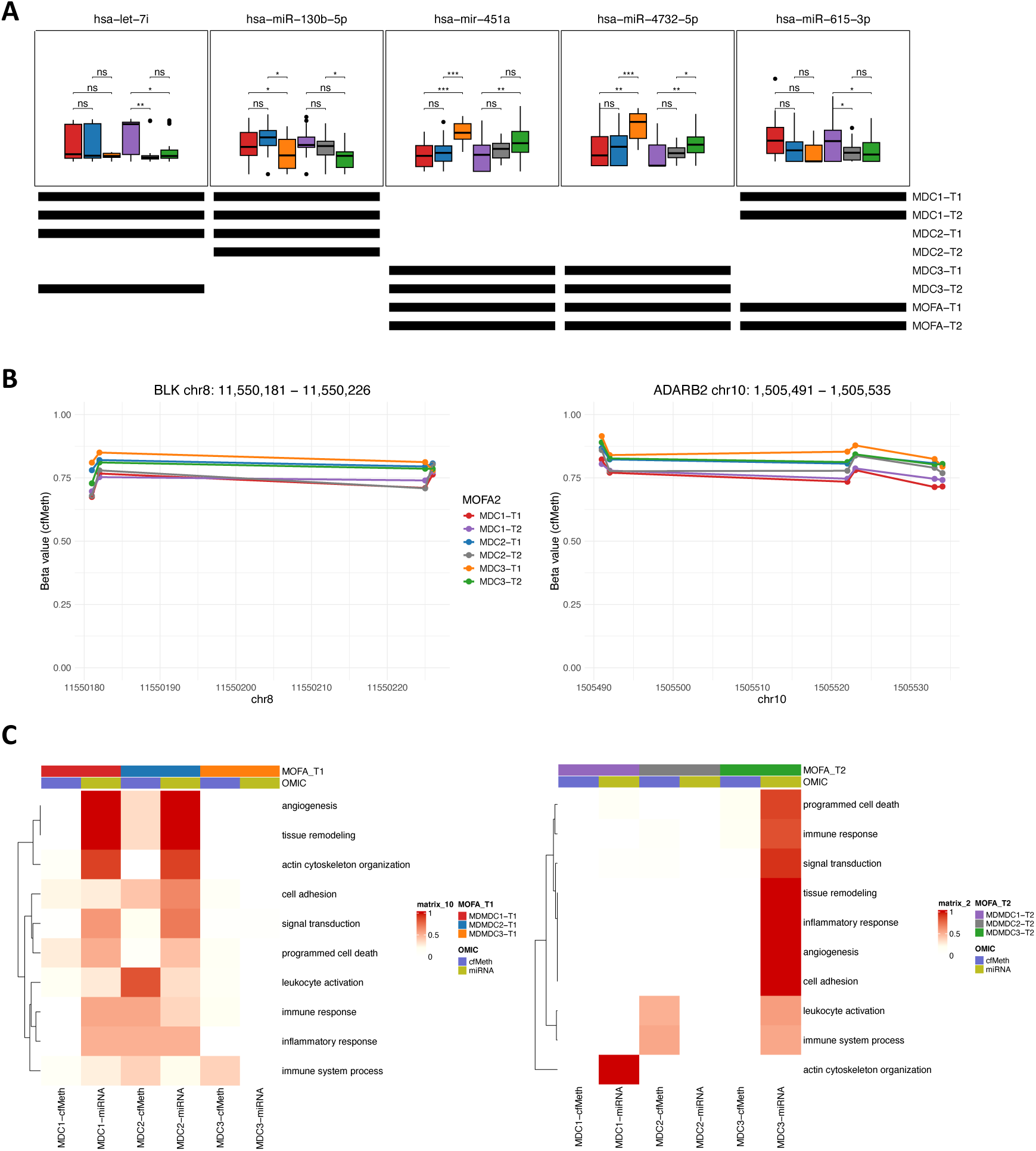
Key molecular features and functional GO analysis underlying MDC subtypes across single-omic analysis. (A) Expression profiles of the five most consistently enriched EVs miRNAs across both single-omic differential expression and MOFA2 integrative models at T1 and T2. Boxplots show normalized expression across MDC subtypes, with heatmap annotation summarizing in which analyses each miRNA showed significance. (B) Methylation levels of two consistently differentially methylated regions annotated to *BLK* and *ADARB2,* across MDC subtypes. (C) Functional enrichment heatmap showing GO pathway for differentially expressed miRNAs and differentially methylated regions across MDC-T1 and MDC-T2. Each cell represents the total number of significantly enriched processes (adjusted p-value < 0.05) aggregated under ten major functional categories (rows) at each omics layers and MDC subtype (columns).

Collectively, these analyses delineate distinct molecular signatures—both transcriptomic and epigenetic—that define the MDC subtypes at baseline and during early treatment. These molecular stratifications not only capture the heterogeneity of NSCLC under ICI but also suggest divergent biological programs that may underlie differential clinical outcomes.

To further dissect the functional relevance of each MDC, we performed pathway enrichment analyses on both omic layers—miRNA and cfDNA methylation—for each molecular subtype. The analyses were constrained to ten major biological themes of relevance to cancer progression and immune dynamics: angiogenesis, tissue remodeling, actin cytoskeleton organization, cell adhesion, signal transduction, programmed cell death, leukocyte activation, immune response, inflammatory response, and immune system process. Although the functional analysis was restricted to a high-level overview of 10 without a detailed assessment of pathway activation or inhibition, the enrichment of specific pathways in individual subtypes aligns with their prognostic profiles. The relative enrichment of each GO category is summarized in Figure 6-C, highlighting both the diversity and specificity of biological processes active within each MDC.

In the T1 model, MDC1-T1—the subtype associated with the poorest overall and progression-free survival—exhibited widespread activation of all these pathways through miRNA expression. Notably, strong enrichment was observed in categories related to angiogenesis, tissue remodeling, and cytoskeletal dynamics, suggesting an aggressive and invasive tumor phenotype. In contrast, cfDNA methylation data contributed minimally to pathway activity in this subtype, pointing to a miRNA regulated biology. MDC2-T1, which showed intermediate survival outcomes, presented a pathway activation profile that partially mirrored MDC1-T1 at the miRNA level. However, unlike MDC1-T1, this subtype also demonstrated marked epigenetic activation— through cfMeth features—of immune-related pathways, including leukocyte activation, immune response, inflammatory response, and immune system process. This dual activation pattern suggests a tumor microenvironment shaped by both oncogenic and immune regulatory signals. Strikingly, MDC3-T1—the group with the most favorable clinical outcomes—displayed minimal pathway activation across both omic layers. A limited enrichment of immune system process was observed in cfMeth data.

In the T2 model, MDC3-T2-corresponding to the most favorable prognosis—showed strong and coordinated activation of nearly all functional exhibited pathways, particularly via miRNA expression, suggesting a complex yet regulated immune and tissue remodeling response under treatment. This widespread activity likely reflects adaptive immune surveillance or a favorable treatment-induced remodeling state. Conversely, MDC1-T2 showed highly selective activation, limited to actin cytoskeleton organization through miRNAs, potentially indicative of ongoing tumor invasiveness or resistance mechanisms. MDC2-T2, in turn, was characterized by focused cfMeth-driven enrichment in immune-related categories, particularly leukocyte activation and immune system process. Although through seemingly different mechanisms, these both subtypes exhibited reduced overall survival.

In summary, transcriptomic and epigenetic profiling reveal that each MDC subtype is driven by distinct biological programs, with varying degrees of immune engagement and tumor-intrinsic activity. These differences may underlie the divergent clinical outcomes observed across clusters. To further explore the immune contexture of each subtype, we next examined peripheral immune cell populations in the same cohort through flow cytometry profiling.

### Immune phenotyping by Flow Cytometry and clinical characterization across MDC subtypes

Beyond molecular characterization, we sought to evaluate whether the MDC subtypes exhibited differential systemic immune phenotypes by analyzing the PBMCs of the same patients using flow cytometry analysis (Figure 7) (Supplementary Table S5). In the T1 model (Figure 7-A), significant differences in T cell populations were observed across MDCs. MDC3-T1 showed reduced frequencies of CD4⁺ T cells and regulatory T cells along with a notable enrichment in cytotoxic CD8⁺ T cells. These findings suggest that the improved clinical outcomes in MDC3-T1 may be linked to a cytotoxic-dominant immune profile and reduced immunosuppression in circulation. In the T2 model (Figure 7-B), CD4⁺ T cell frequencies were again significantly altered, this time higher in MDC3-T2 compared to MDC1-T2. Moreover, exhausted CD8+ T cells– CD8⁺PD1⁺ T cells– were differentially distributed across subtypes: the abundance was highest in MDC1-T2, intermediate in MDC3-T2, and lowest in MDC2-T2. In contrast, CD3⁺PD-L1⁺ T cells showed an inverse distribution, being most frequent in MDC2-T2 and least in MDC1-T2. Additionally, CD19⁺ B cells differed between subtypes, with MDC2-T2 displaying elevated frequencies relative to MDC1-T2. Together, these immune profiling data indicate that each MDC subtype is not only defined by distinct molecular signatures but is also accompanied by unique circulating immune landscapes.

**Figure 7.**
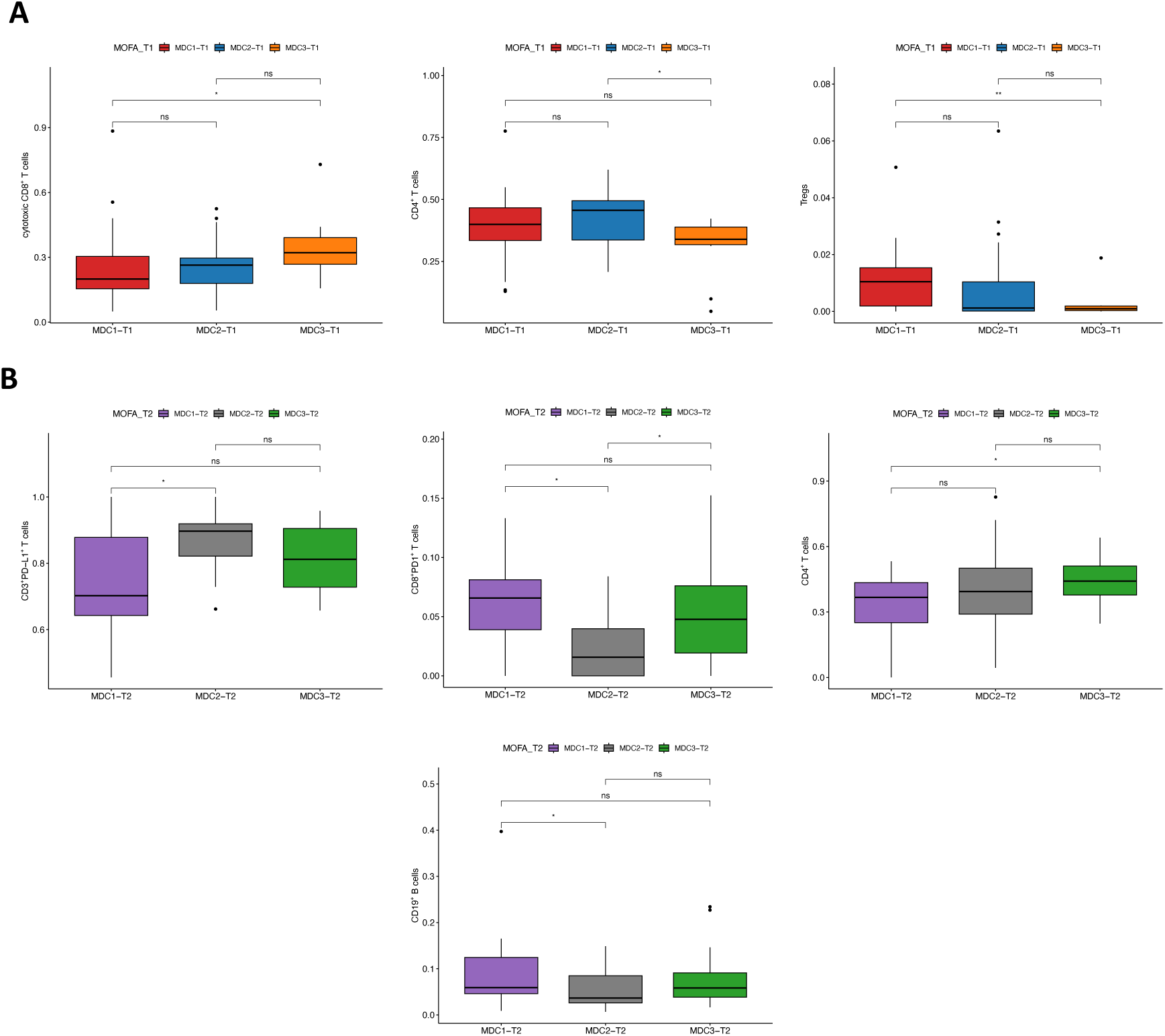
Immune phenotyping by flow cytometry across MDC subtypes. (A) Immune cell composition across MDC-T1 subtypes. (B) Immune phenotyping across MDC-T2 subtypes. Boxplots illustrate the relative abundance of selected immune populations measured by flow cytometry. Y-axis values represent the percentage of each immune population relative to its parent gate. Comparisons between MDC subtypes were assessed using pairwise Wilcoxon rank-sum tests, with p-values indicated within each panel. Significance is denoted as ***p < 0.001; **p < 0.01; *p < 0.05; ns: not significant.

In addition to their molecular stratification, MOFA2 latent factors and their corresponding MDCs demonstrated significant associations with clinical variables (Supplementary Figure S6). In the T1 model, all three latent factors were significantly elevated in patients exhibiting primary resistance to ICI therapy, suggesting that early molecular programs captured by MOFA2 reflect intrinsic mechanisms of treatment failure. F2 was also significantly higher in patients with PD-L1 expression TPS <50%, further supporting its link to immune escape phenotypes. At the cluster level, MDC3-T1, characterized by favorable survival, was enriched for non-resistant patients, while cutaneous metastases were less frequent in MDC1-T1 and MDC2-T1, indicating distinct patterns of disease dissemination among subtypes.

In the T2 model, a similar clinical alignment was observed. F1 was associated with primary resistance, whereas F2 and F3 were significantly enriched in non-resistant patients, mirroring the survival hierarchy observed across MDCs. F5 was inversely associated with cutaneous metastases, suggesting this latent component may capture systemic progression phenotypes. Consistent with earlier findings, MDC3-T2, the most favorable subtype, was predominantly composed of non-resistant cases, reinforcing the clinical validity of MOFA2-derived subtypes over time.

### External validation confirms the clinical relevance of MDC subtypes

Given the lack of publicly available datasets profiling both cfDNA methylation and EV-derived miRNAs in the same ICI-treated NSCLC patients, we conducted external validation independently for each omic layer. To this end, MDC signatures from T1 and T2 were validated using baseline miRNA expression data from the Genova et al., 2024 cohort and tumor methylation profiles from the SMC cohort, respectively. For each omic and timepoint, we developed predictive models to assign MDC labels in the external samples, using only features overlapping with those in our dataset. Models were trained using supervised learning algorithms—including Lasso logistic regression, SVMRadial, and RF classifiers—and were evaluated by cross-validation to identify the approach yielding the highest generalizability based on AUC (Supplementary Table S6). Once MDC labels were assigned, survival analyses were performed for each validation cohort.

In the cfDNA methylation validation using the SMC dataset (Figure 8-A), the MDC-T1 model yielded a PFS stratification trend approaching statistical significance (log-rank p = 0.065), reproducing the same prognostic pattern observed in our cohort: MDC1-T1 associated with poor outcome, MDC2-T1 intermediate, and MDC3-T1 favorable. More robust results were obtained for MDC-T2, where significant differences in PFS were observed (log-rank p = 0.026), with MDC3-T2 showing the longest median PFS. In the miRNA validation using the Genova cohort, the MDC-T1 model failed to assign any samples to the MDC3-T1 group (Figure 8-B), and survival differences were not statistically significant for either OS or PFS. However, in the MDC-T2 model (Figure 8-C), the subtype MDC3-T2 again emerged as a best-prognosis group, showing significant survival differences with a median OS and PFS of log-rank p = 0.047 and 0.0038, respectively.

**Figure 8.**
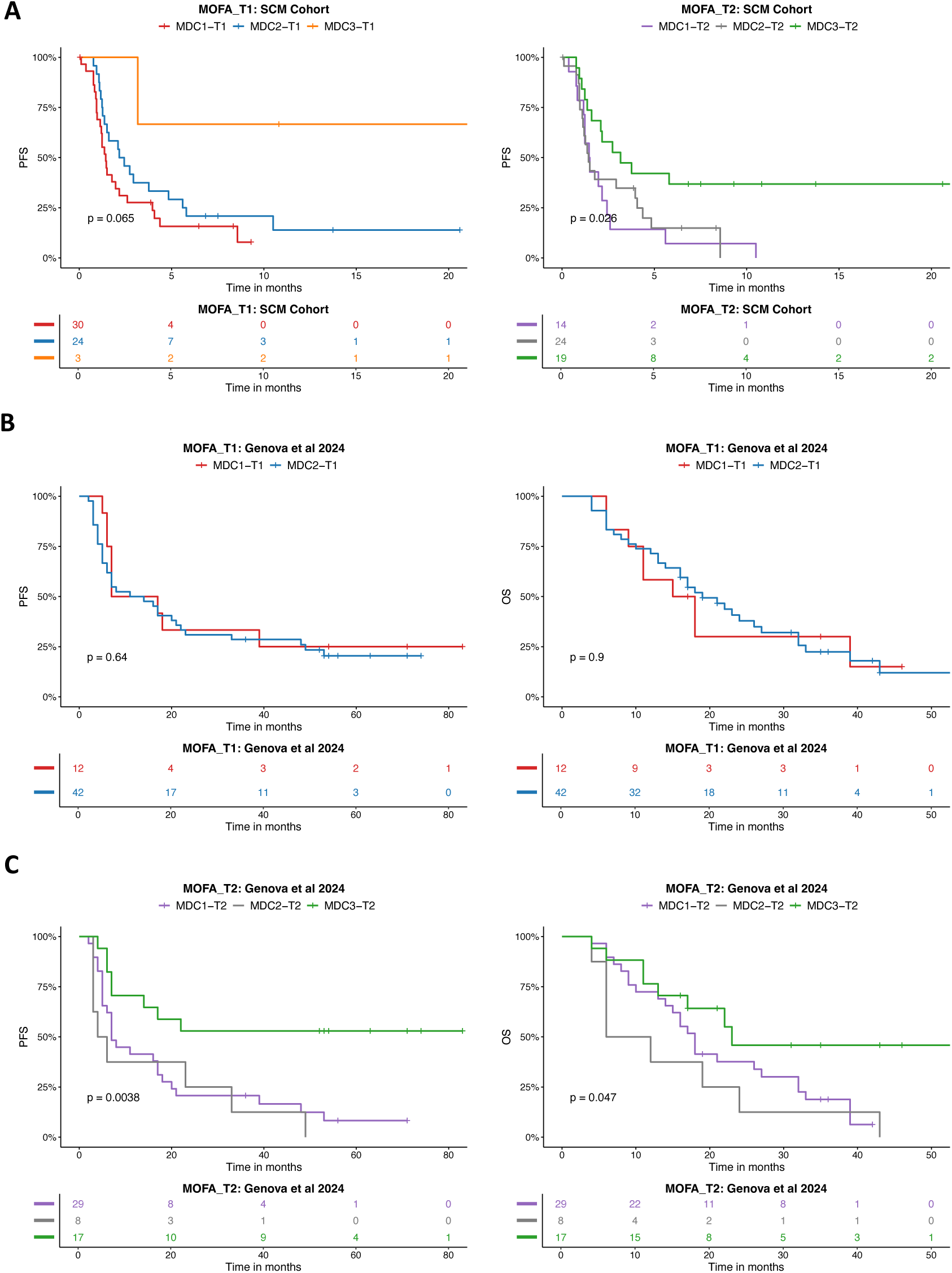
External validation of MDCs in independent cfMeth and miRNA cohorts. (A) Kaplan– Meier curves of PFS for MDC-T1 and MDC-T2 subtypes predicted onto the SMC dataset (tumor tissue methylation, n=57). (B) OS and PFS stratification based on MDC-T1 subtype predictions in the Genova et al., 2024 cohort (EV-derived miRNA, n=54). (C) OS and PFS stratification based on MDC-T2 subtype predictions in the Genova et al., 2024 cohort. Kaplan–Meier curves show survival distributions where p-values were calculated using the log-rank test.

Together, these results demonstrate a robust multi-omic landscape of immune checkpoint blockade in NSCLC, identifying reproducible molecular subtypes through integrative profiling of distinct circulating biomarkers.

## Discussion

Non-small cell lung cancer exhibits inter-patient heterogeneity in response to ICIs, and recent studies underscore the urgent need for robust biomarkers to guide treatment decisions ^3,7^. Leveraging a longitudinal design and a multi-omics integration strategy based on MOFA2, our study identifies molecular subtypes — named MDCs—at two key treatment stages: baseline (T1) and early on-treatment (T2). These subtypes are not only distinct in their molecular architecture but also associated with significantly different survival outcomes. By integrating cfDNA methylation and extracellular vesicle-derived miRNA profiles, we demonstrate that MDCs represent biologically meaningful clusters, offering a fine-grained molecular stratification framework for ICI-treated NSCLC patients.

Previous studies have demonstrated the utility of unsupervised multi-omic approaches to dissect patient heterogeneity and uncover latent biological patterns associated with clinical outcomes, particularly in the context of immune-based therapies ^28,29^. While integrative analyses using bulk tumor tissue have provided valuable insights into responder versus non-responder classification in NSCLC ^30,31^, our study takes a distinct approach by leveraging a liquid biopsy-based strategy and applying factor analysis within an unsupervised framework. This methodology enables the identification of intrinsic molecular subtypes without prior outcome labeling, thus reducing bias and enhancing biological interpretability ^32,33^. Moreover, we focus on the integration of two interrelated liquid biopsy omics—cfDNA methylation and EV-derived miRNAs—which have individually demonstrated prognostic value in NSCLC ^34–37^. The biological complementarity of these two omic layers is well established, with evidence showing mechanistic links between miRNA expression and DNA methylation regulation ^38,39^. While cfDNA methylation offers a window into epigenetic remodeling and immune landscape ^40^, EV-miRNAs have been extensively studied in ICI-treated NSCLC as both prognostic and predictive biomarkers ^26,41,42^. Collectively, this integrative, non-invasive framework not only enhances molecular resolution but also offers translational potential for future biomarker development in NSCLC.

At baseline (T1), MDC1-T1 emerged as the molecular subtype with the poorest OS and PFS. This cluster was defined by high MOFA2 scores in Factors 1 and 3, with *hsa-miR-200c-3p* standing out as a key molecular driver. Interestingly, previous research in epithelial ovarian cancer suggests that *hsa-miR-200c-3p* concomitantly reduced PD-L1 expression ^43^. This finding may reflect regulatory effects or feedback mechanisms, whereby increased *hsa-miR-200c-3p* coincides with reduced PD-L1 levels, potentially promoting ICI resistance. In our context, this miRNA was markedly overexpressed in MDC1-T1 and associated with adverse clinical outcomes. Pathway enrichment analyses revealed that MDC1-T1 was primarily driven by miRNA signatures activating angiogenesis, tissue remodeling, and cytoskeletal dynamics—hallmarks of an aggressive, invasive tumor phenotype. Moreover, cfDNA methylation (cfMeth) data contributed minimally to pathway activity in this subtype, indicating a lack of epigenetically driven signaling and suggesting a biology primarily orchestrated at transcriptomic layer. Conversely, MDC2-T1 presented with intermediate survival and a dual-layered activation pattern, encompassing both miRNA and cfMeth features. Notably, this subtype showed upregulation of *hsa-miR-133a-3p*, a known tumor suppressor that induces apoptosis and suppresses proliferation via the EGFR/AKT/ERK axis in NSCLC cells ^44^. Furthermore, epigenetic dysregulation of *BAP1* was observed in MDC2-T1, suggesting elevated tumor expression levels in this subtype. In malignant pleural mesothelioma, *BAP1*^low^ tumors have been shown to reside in close proximity to T cells, potentially enhancing responsiveness to ICI therapy ^45^, hinting at a complex immunomodulatory role. This interplay of oncogenic suppression and immune evasion may partially explain the intermediate prognosis observed in this group. MDC3-T1, by contrast, was consistently associated with favorable outcomes and displayed minimal activation across both transcriptomic and epigenetic layers, suggesting a quiescent state compare with the other subtypes in MDC-T1. Two miRNAs—*hsa-miR-451a* and *hsa-miR-4732-5p*—emerged as prominent features within this cluster. *hsa-miR-451a*, in particular, has been implicated as a circulating biomarker for immune regulation and has been shown to be enriched in extracellular vesicles ^46^. High levels of *hsa-miR-451a* have also been associated with improved OS in lung squamous cell carcinoma ^47^, supporting its protective role and its potential as a minimally invasive prognostic marker. In addition, *hsa-miR-4732-5p* overexpression has been shown to suppress proliferation, migration, and invasion of NSCLC cells ^48^. Altogether, the transcriptional and epigenetic landscapes of the T1-derived subtypes provide mechanistic insight into the biological diversity of NSCLC and its influence on early ICI response.

At the early on-treatment time point (T2), the integrative molecular landscape revealed different subtypes that only partially overlapped with those identified at baseline (MDC-T1), highlighting the dynamic nature of tumor biology under ICI pressure. MDC3-T2 remained associated with the most favorable prognosis and exhibited robust activation of immune-related and tissue remodeling pathways, predominantly through miRNA-mediated regulation. A key molecular feature of this subtype is *hsa-miR-30a-5p*, which was negatively associated with MOFA Factor 4 and hereby enriched in MDC3-T2. Mechanistically, *hsa-miR-30a-5p* has been implicated in immune evasion, as hsa_circ_0020714 can sponge this miRNA to upregulate *SOX4*, promoting immune escape in NSCLC cells ^49,50^. Additionally, MDC3-T2 was linked to a hypomethylated region within the *FBXW7* locus, suggesting increased gene expression. Notably, higher *FBXW7* levels have been shown to attenuate drug resistance and enhance the efficacy of immunotherapy, correlating with improved survival outcomes in NSCLC ^51^. In contrast, MDC1-T2 was characterized by selective activation of actin cytoskeleton organization pathways via miRNA expression, a signature associated with increased cellular motility and immune evasion ^52^. MDC2-T2, meanwhile, exhibited prominent cfDNA methylation-driven enrichment of immune-related pathways, potentially indicative of an epigenetically encoded immune-resistant phenotype.

Interestingly, both MDC1-T2 and MDC2-T2 shared similar overall and progression-free survival profiles, which may be partially explained by shared molecular drivers such as *hsa-miR-106b-5p*. This miRNA was upregulated in both subtypes and is known to promote NSCLC progression by downregulating *BTG3*, a tumor suppressor involved in cell cycle control ^53^.

Moreover, beyond the key features previously discussed, additional molecules further underscore the biological coherence across MDC subtypes. Notably, *hsa-let-7i-5p* and *hsa-miR-130b-5p* emerged as recurrent markers in poor-outcome subtypes such as MDC1-T1, MDC2-T1, and MDC1-T2. These miRNAs have been reported to exhibit oncogenic properties of tumor promoting in NSCLC ^54^ and in other malignancies ^55^. At the epigenetic level, two differentially methylated regions located within the genes *BLK* and *ADARB2* were consistently hypermethylated in subtypes associated with the most favorable survival outcomes—MDC3-T1 and MDC3-T2. This hypermethylation likely contributes to downregulation of *BLK*, aligning with prior studies that demonstrated high *BLK* expression promotes tumor growth and malignant transformation in B-cell lymphoma models ^56^. While no direct literature links *ADARB2* to immunotherapy response, its paralog *ADAR1* has been implicated in resistance to ICIs. Loss of *ADAR1* was shown to overcome immune evasion in tumors with defective antigen presentation, thus restoring sensitivity to PD-1 blockade ^57^. This raises the possibility that epigenetic regulation of *ADARB2* may reflect broader RNA editing–related mechanisms influencing immune response in NSCLC, warranting further investigation.

The clinical relevance of the identified MDC subtypes was further underscored by their association with primary resistance to ICI therapy. In both T1 and T2 models, resistance was predominantly grouped to subtypes with poorer clinical outcomes—namely, MDC1 and MDC2— while non-responder patients were largely enriched within MDC3-T1 and MDC3-T2. Additional clinical stratification revealed that Factor 2 in the T1 model was significantly elevated in patients with PD-L1 TPS below 50%, a subgroup previously associated with reduced response rates and worse outcomes to ICI therapies ^4^. Cutaneous metastases were also unevenly distributed across MDC-T1 subtypes; however, given the limited literature available on this variable in the context of ICI-treated NSCLC, further investigation is warranted to elucidate its potential biological implications. Complementing the molecular stratification, immunophenotyping by flow cytometry revealed distinct immune cell profiles across MDCs. MDC3-T1, which showed the most favorable prognosis, was enriched in circulating cytotoxic CD8⁺ T cells and displayed a relative depletion of regulatory T cells, a pattern indicative of an effective cytotoxic immune landscape. In contrast, MDC2-T1 exhibited elevated levels of CD4⁺ T cells, which, although less characterized, have been reported as correlating with clinical benefit and response to ICIs in NSCLC ^58^. These findings align with prior studies highlighting the prognostic and predictive relevance of CD8⁺ T cells both in tumor tissue ^59^ and peripheral blood ^60^, as well as the association of baseline Treg depletion with improved ICI outcomes ^61^. By the T2 timepoint, immune profiles further diverged. MDC3-T2 was characterized by increased CD4⁺ T cells, while MDC2-T2 demonstrated a marked depletion of exhausted T cells (CD8⁺PD1⁺ T cells)—a population previously associated with durable responses and prolonged PFS in ICI-treated NSCLC patients^62^. Additionally, this subtype showed increased levels of CD3⁺PD-L1⁺ T cells, a potentially adaptive mechanism reflecting impaired cytotoxic T cell activity or a shift toward immune exhaustion. Collectively, these results bridge the molecular and immunological dimensions of MDC stratification, reinforcing their potential as clinically actionable biomarkers for immunotherapy in NSCLC.

Persistence and prognostic utility across treatment timepoints were assessed through the projection of MOFA-derived molecular subtypes onto longitudinally collected samples (T1–T4). Notably, the MDC-T2 model demonstrated superior temporal generalizability, maintaining consistent prognostic stratification in both earlier and later samples. This suggests that the T2-derived molecular programs may better capture treatment-modulated biological states than those defined at baseline. Crucially, the robustness of these subtypes was validated in independent external cohorts. Despite differences in sample type, platform, and omic layer, the cfMeth-based MDC-T2 model significantly stratified PFS in the SMC dataset ^27^. Likewise, MDC2-T2 consistently predicted both OS and PFS in the Genova et al. ^26^ EVs-miRNA cohort. These findings underscore the reproducibility and potential clinical transferability of the MDC classification.

Nevertheless, several methodological limitations should be taken into consideration. The absence of external datasets with paired multi-omic liquid biopsy profiles constrained validation to single-omic projections, thereby limiting the holistic evaluation of integrated signatures. For cfDNA methylation, validation was performed on tumor tissue samples rather than plasma-derived cfDNA, introducing biological discrepancies in the origin and context of methylation signals. Similarly, miRNA validation relied on microarray data, which only partially overlaps with the small RNA sequencing data used in our study cohort, potentially reducing marker concordance. These discrepancies highlight a critical need for harmonized, multi-modal profiling platforms in future prospective studies to enable more accurate translational implementation. Moreover, Further comparative studies across different malignancies and treatment settings— beyond ICI—as well as investigations into the profile and impact of immune-related toxicities, will provide valuable insights into the relevance and broader applicability of the proposed subtypes.

In summary, our study presents a comprehensive and clinically actionable framework for molecular subtyping of metastatic NSCLC based on integrated liquid biopsy-derived miRNA and cfDNA methylation profiles. The identification of MDCs at both baseline and early on-treatment timepoints provides meaningful prognostic information and captures dynamic, treatment-influenced biology. Importantly, individual omic or immunological correlates alone are insufficient to fully capture the complexity of the mechanisms underlying the proposed subtypes, underscoring the need for integrated multi-omic analysis. These findings support the incorporation of multi-layered molecular profiling into the future of precision immuno-oncology and lay the groundwork for biomarker-driven, personalized therapeutic strategies in NSCLC.

## Conclusion

Overall, this study demonstrates that integrative multi-omic profiling of cfDNA methylation and EV-derived miRNAs from liquid biopsies can effectively capture the molecular heterogeneity of NSCLC patients undergoing immunotherapy. Through unsupervised modeling, we identified distinct MDC subtypes associated with divergent clinical outcomes and immune phenotypes, which were validated across longitudinal samples and external datasets. These subtypes reflect biologically coherent programs relevant to ICI response, such as angiogenesis, immune modulation, and cytoskeletal remodeling. Importantly, the multi-omic MDC framework provides a more refined stratification offering novel insights into tumor-immune dynamics. Our findings underscore the utility of liquid biopsy-based, multi-omic approaches for precision immuno-oncology and advocate for their integration in future prospective trials to optimize therapeutic decision-making in NSCLC.

## Declarations

### Ethics approval and consent to participate

The study follows the Declaration of Helsinki and has been submitted and approved by Comité de Ética de la Investigación Provincial de Málaga. Approval date on 26 October 2017 with the title: “Omics integration for precision cancer immunotherapy” (799818, H2020-MSCA-IF-2017) research project. All patients signed Informed Consent to participate in the study and received an information sheet about the project.

## Consent for publication

Not applicable.

## Data availability

The original contributions presented in the study are included in the article Supplementary Material and Supplementary Tables, further inquiries can be directed to the corresponding author. Data is available upon reasonable request. All data analyzed in our study are available upon request.

## Code availability

Code available upon request on http://github.com/ImmunoOncology.

## Author contributions

**J.L. O.**: Conceptualization, design of the work, interpretation of data, investigation, formal analysis, data curation, and writing – original draft. **E. P.-R.**: Conceptualization, data acquisition, interpretation of data, and review. **L. F.-O., and B. M.-G.**: Formal analysis, laboratory experiments and review. **J.M. J. and J.C. B.**: Data acquisition and review. **A. R.-D.**: Investigation, design of the work, data acquisition, supervision, and writing – review and editing. **I. B.**: Conceptualization, design of the work, interpretation of data, supervision, and writing – review and editing. All authors reviewed and approved the final manuscript.

## Data Availability

The original contributions presented in the study are included in the article Supplementary Material and Supplementary Tables, further inquiries can be directed to the corresponding author. Data is available upon reasonable request. All data analyzed in our study are available upon request. Code available upon request on http://github.com/ImmunoOncology

## Acknowledgments

The authors gratefully acknowledge the Supercomputing and Bioinnovation Center (SCBI) at the University of Malaga for providing computational resources (http://www.scbi.uma.es/). They are also grateful to the Hospital-IBIMA Biobank (Biobanco del Sistema Sanitario Público de Andalucía) integrated in the Spanish National biobanks Network (PT23/00049) supported by ISCIII. Additionally, the authors express their gratitude to Andrea González, Marina Rivero, María Garrido, Javier Oliver and Alberto Ríos for their valuable support and insightful suggestions, which significantly enhanced the quality of this work.

## List of abbreviations

5hmC: 5-hydroxymethylcytosine
5mC: 5-methylcytosine
AUC: Area Under the Curve
cfDNA: Circulating Free DNA
cfMeth: Circulating cell-free DNA Methylation
CTCs: Circulating Tumor Cells
ECOG: Eastern Cooperative Oncology Group
EM-seq: Enzymatic Methyl-sequencing
EVs: Extracellular Vesicles
FDA: Food and Drug Administration
ICIs: Immune Checkpoint Inhibitors
irAEs: Immune-related Adverse Events
IT: Immunotherapy
LUAD: Lung Adenocarcinoma
LUSC: Lung Squamous Cell Carcinoma
MDC: MOFA2 derived cluster
miRNAs: MicroRNAs
MOFA2: Multi-Omics Factor Analysis version 2
NCCN: National Comprehensive Cancer Network
NSCLC: Non-Small Cell Lung Cancer
OS: Overall Survival
PAM: Partitioning Around Medoids
PBMCs: Peripheral Blood Mononuclear Cells
PFS: Progression-Free Survival
RF: Random Forest
SVM-Radial: Support Vector Machine with Radial Kernel
TMB: Tumor Mutational Burden
TPS: Tumor Proportion Score
Tregs: Regulatory T Cells

## References

1. Bray F, Laversanne M, Sung H, et al. Global cancer statistics 2022: GLOBOCAN estimates of incidence and mortality worldwide for 36 cancers in 185 countries. CA Cancer J Clin. 2024;74(3):229-263. doi:10.3322/caac.21834

2. Herbst RS, Morgensztern D, Boshoff C. The biology and management of non-small cell lung cancer. Nature. 2018;553(7689):446-454. doi:10.1038/nature25183

3. Cheng Y, Zhang T, Xu Q. Therapeutic advances in non-small cell lung cancer: Focus on clinical development of targeted therapy and immunotherapy. MedComm. 2021;2(4):692–729. doi:10.1002/mco2.105

4. Mok TSK, Wu YL, Kudaba I, et al. Pembrolizumab versus chemotherapy for previously untreated, PD-L1-expressing, locally advanced or metastatic non-small-cell lung cancer (KEYNOTE-042): a randomised, open-label, controlled, phase 3 trial. Lancet Lond Engl. 2019;393(10183):1819-1830. doi:10.1016/S0140-6736(18)32409-7

5. Giroux Leprieur E, Dumenil C, Julie C, et al. Immunotherapy revolutionises non-small-cell lung cancer therapy: Results, perspectives and new challenges. Eur J Cancer. 2017;78:16–23. doi:10.1016/j.ejca.2016.12.041

6. Reck M, Rodríguez-Abreu D, Robinson AG, et al. Pembrolizumab versus Chemotherapy for PD-L1–Positive Non–Small-Cell Lung Cancer. N Engl J Med. 2016;375(19):1823–1833. doi:10.1056/NEJMoa1606774

7. Suresh K, Naidoo J, Lin CT, Danoff S. Immune Checkpoint Immunotherapy for Non-Small Cell Lung Cancer: Benefits and Pulmonary Toxicities. Chest. 2018;154(6):1416–1423. doi:10.1016/j.chest.2018.08.1048

8. Davis AA, Patel VG. The role of PD-L1 expression as a predictive biomarker: an analysis of all US Food and Drug Administration (FDA) approvals of immune checkpoint inhibitors. J Immunother Cancer. 2019;7(1):278. doi:10.1186/s40425-019-0768-9

9. Vaddepally RK, Kharel P, Pandey R, Garje R, Chandra AB. Review of Indications of FDA-Approved Immune Checkpoint Inhibitors per NCCN Guidelines with the Level of Evidence. Cancers. 2020;12(3):738. doi:10.3390/cancers12030738

10. Nicoś M, Krawczyk P, Crosetto N, Milanowski J. The Role of Intratumor Heterogeneity in the Response of Metastatic Non-Small Cell Lung Cancer to Immune Checkpoint Inhibitors. Front Oncol. 2020;10:569202. doi:10.3389/fonc.2020.569202

11. Marabelle A, Fakih M, Lopez J, et al. Association of tumour mutational burden with outcomes in patients with advanced solid tumours treated with pembrolizumab: prospective biomarker analysis of the multicohort, open-label, phase 2 KEYNOTE-158 study. Lancet Oncol. 2020;21(10):1353–1365. doi:10.1016/S1470-2045(20)30445-9

12. Yang Y, Liu H, Chen Y, et al. Liquid biopsy on the horizon in immunotherapy of non-small cell lung cancer: current status, challenges, and perspectives. Cell Death Dis. 2023;14(3):230. doi:10.1038/s41419-023-05757-5

13. Karachaliou N, Mayo-de-las-Casas C, Molina-Vila MA, Rosell R. Real-time liquid biopsies become a reality in cancer treatment. Ann Transl Med. 2015;3(3):36. doi:10.3978/j.issn.2305-5839.2015.01.16

14. Fabbri M, Calin GA. 4 - Epigenetics and miRNAs in Human Cancer. In: Herceg Z, Ushijima T, eds. Advances in Genetics. Vol 70. Epigenetics and Cancer, Part A. Academic Press; 2010:87–99. doi:10.1016/B978-0-12-380866-0.60004-6

15. Kluger HM, Tawbi HA, Ascierto ML, et al. Defining tumor resistance to PD-1 pathway blockade: recommendations from the first meeting of the SITC Immunotherapy Resistance Taskforce. J Immunother Cancer. 2020;8(1):e000398. doi:10.1136/jitc-2019-000398

16. Vaisvila R, Ponnaluri VKC, Sun Z, et al. Enzymatic methyl sequencing detects DNA methylation at single-base resolution from picograms of DNA. Genome Res. 2021;31(7):1280–1289. doi:10.1101/gr.266551.120

17. Onieva JL, Xiao Q, Berciano-Guerrero MÁ, et al. High IGKC-Expressing Intratumoral Plasma Cells Predict Response to Immune Checkpoint Blockade. Int J Mol Sci. 2022;23(16):9124. doi:10.3390/ijms23169124

18. Krueger F, Andrews SR. Bismark: a flexible aligner and methylation caller for Bisulfite-Seq applications. Bioinforma Oxf Engl. 2011;27(11):1571–1572. doi:10.1093/bioinformatics/btr167

19. Li J, Kho AT, Chase RP, et al. COMPSRA: a COMprehensive Platform for Small RNA-Seq data Analysis. Sci Rep. 2020;10(1):4552. doi:10.1038/s41598-020-61495-0

20. Love MI, Huber W, Anders S. Moderated estimation of fold change and dispersion for RNA-seq data with DESeq2. Genome Biol. 2014;15(12):550. doi:10.1186/s13059-014-0550-8

21. Kern F, Fehlmann T, Solomon J, et al. miEAA 2.0: integrating multi-species microRNA enrichment analysis and workflow management systems. Nucleic Acids Res. 2020;48(W1):W521–W528. doi:10.1093/nar/gkaa309

22. Akalin A, Kormaksson M, Li S, et al. methylKit: a comprehensive R package for the analysis of genome-wide DNA methylation profiles. Genome Biol. 2012;13(10):R87. doi:10.1186/gb-2012-13-10-r87

23. Yu G, Wang LG, Han Y, He QY. clusterProfiler: an R Package for Comparing Biological Themes Among Gene Clusters. OMICS J Integr Biol. 2012;16(5):284–287. doi:10.1089/omi.2011.0118

24. Argelaguet R, Arnol D, Bredikhin D, et al. MOFA+: a statistical framework for comprehensive integration of multi-modal single-cell data. Genome Biol. 2020;21(1):111. doi:10.1186/s13059-020-02015-1

25. Onieva JL, Cháves P, Oliver J, et al. Abstract 1910: flowTOTAL: A comprehensive bioinformatics workflow for flow cytometry automatic analysis. Cancer Res. 2022;82(12_Supplement):1910. doi:10.1158/1538-7445.AM2022-1910

26. Genova C, Marconi S, Chiorino G, et al. Extracellular vesicles miR-574-5p and miR-181a-5p as prognostic markers in NSCLC patients treated with nivolumab. Clin Exp Med. 2024;24(1):182. doi:10.1007/s10238-024-01427-8

27. Kim K, Kim H, Shin I, et al. Genomic hypomethylation in cell-free DNA predicts responses to checkpoint blockade in lung and breast cancer. Sci Rep. 2023;13(1):22482. doi:10.1038/s41598-023-49639-4

28. Pekayvaz K, Losert C, Knottenberg V, et al. Multiomic analyses uncover immunological signatures in acute and chronic coronary syndromes. Nat Med. 2024;30(6):1696–1710. doi:10.1038/s41591-024-02953-4

29. Sharma A, Debik J, Naume B, Ohnstad HO, Bathen TF, Giskeødegård GF. Comprehensive multi-omics analysis of breast cancer reveals distinct long-term prognostic subtypes. Oncogenesis. 2024;13(1):1–13. doi:10.1038/s41389-024-00521-6

30. Yan Y, Sun D, Hu J, et al. Multi-omic profiling highlights factors associated with resistance to immuno-chemotherapy in non-small-cell lung cancer. Nat Genet. 2025;57(1):126–139. doi:10.1038/s41588-024-01998-y

31. Parra ER, Zhang J, Duose DY, et al. Multi-omics Analysis Reveals Immune Features Associated with Immunotherapy Benefit in Patients with Squamous Cell Lung Cancer from Phase III Lung-MAP S1400I Trial. Clin Cancer Res. 2024;30(8):1655–1668. doi:10.1158/1078-0432.CCR-23-0251

32. Cai Z, Poulos RC, Liu J, Zhong Q. Machine learning for multi-omics data integration in cancer. iScience. 2022;25(2):103798. doi:10.1016/j.isci.2022.103798

33. Argelaguet R, Velten B, Arnol D, et al. Multi-Omics Factor Analysis—a framework for unsupervised integration of multi-omics data sets. Mol Syst Biol. 2018;14(6):e8124. doi:10.15252/msb.20178124

34. Ospina AV. Overview of the Role of Liquid Biopsy in Non-small Cell Lung Cancer (NSCLC). Clin Oncol. 2024;36(10):e371–e380. doi:10.1016/j.clon.2024.07.004

35. Oliver J, Onieva JL, Garrido-Barros M, et al. Fluorometric Quantification of Total Cell-Free DNA as a Prognostic Biomarker in Non-Small-Cell Lung Cancer Patients Treated with Immune Checkpoint Blockade. Cancers. 2023;15(13):3357. doi:10.3390/cancers15133357

36. Janke F, Gasser M, Angeles AK, et al. Low-coverage whole genome sequencing of cell-free DNA to predict and track immunotherapy response in advanced non-small cell lung cancer. J Exp Clin Cancer Res. 2025;44(1):87. doi:10.1186/s13046-025-03348-0

37. Mondelo-Macía P, García-González J, León-Mateos L, et al. Clinical potential of circulating free DNA and circulating tumour cells in patients with metastatic non-small-cell lung cancer treated with pembrolizumab. Mol Oncol. 2021;15(11):2923–2940. doi:10.1002/1878-0261.13094

38. Saviana M, Le P, Micalo L, et al. Crosstalk between miRNAs and DNA Methylation in Cancer. Genes. 2023;14(5):1075. doi:10.3390/genes14051075

39. Saito Y, Saito H, Liang G, Friedman JM. Epigenetic alterations and microRNA misexpression in cancer and autoimmune diseases: a critical review. Clin Rev Allergy Immunol. 2014;47(2):128–135. doi:10.1007/s12016-013-8401-z

40. Luo H, Wei W, Ye Z, Zheng J, Xu R hua. Liquid Biopsy of Methylation Biomarkers in Cell-Free DNA. Trends Mol Med. 2021;27(5):482–500. doi:10.1016/j.molmed.2020.12.011

41. Peng XX, Yu R, Wu X, et al. Correlation of plasma exosomal microRNAs with the efficacy of immunotherapy in EGFR/ALK wild-type advanced non-small cell lung cancer. J Immunother Cancer. 2020;8(1):e000376. doi:10.1136/jitc-2019-000376

42. Abdipourbozorgbaghi M, Vancura A, Radpour R, Haefliger S. Circulating miRNA panels as a novel non-invasive diagnostic, prognostic, and potential predictive biomarkers in non-small cell lung cancer (NSCLC). Br J Cancer. 2024;131(8):1350–1362. doi:10.1038/s41416-024-02831-3

43. Anastasiadou E, Messina E, Sanavia T, et al. MiR-200c-3p Contrasts PD-L1 Induction by Combinatorial Therapies and Slows Proliferation of Epithelial Ovarian Cancer through Downregulation of β-Catenin and c-Myc. Cells. 2021;10(3):519. doi:10.3390/cells10030519

44. Guo N, Zhao Y, Zhang W, Li S, Li S, Yu J. MicroRNA-133a downregulated EGFR expression in human non-small cell lung cancer cells via AKT/ERK signaling. Oncol Lett. 2018;16(5):6045–6050. doi:10.3892/ol.2018.9399

45. Xu D, Gao Y, Yang H, et al. BAP1 Deficiency Inflames the Tumor Immune Microenvironment and Is a Candidate Biomarker for Immunotherapy Response in Malignant Pleural Mesothelioma. JTO Clin Res Rep. 2024;5(5):100672. doi:10.1016/j.jtocrr.2024.100672

46. Wang F xiang, Shi Z an, Mu G. Regulation of immune cells by miR-451 and its potential as a biomarker in immune-related disorders: a mini review. Front Immunol. 2024;15. doi:10.3389/fimmu.2024.1421473

47. Gao B, Li R, Song X, Hu S, Yang F. miR-139-5p and miR-451a as a Diagnostic Biomarker in LUSC. Pharmacogenomics Pers Med. 2023;16:313–323. doi:10.2147/PGPM.S402750

48. Tang X, Liu S, Cui Y, Zhao Y. MicroRNA-4732 is downregulated in non-small cell lung cancer and inhibits tumor cell proliferation, migration, and invasion. Respir Med Res. 2021;80:100865. doi:10.1016/j.resmer.2021.100865

49. Wu J, Zhu MX, Li KS, Peng L, Zhang PF. Circular RNA drives resistance to anti-PD-1 immunotherapy by regulating the miR-30a-5p/SOX4 axis in non-small cell lung cancer. Cancer Drug Resist. 2022;5(2):261–270. doi:10.20517/cdr.2021.100

50. Bagati A, Kumar S, Jiang P, et al. Integrin αvβ6 – TGFβ – SOX4 Pathway Drives Immune Evasion in Triple-negative Breast Cancer. Cancer Cell. 2021;39(1):54–67.e9. doi:10.1016/j.ccell.2020.12.001

51. Chen S, Lin J, Zhao J, et al. FBXW7 attenuates tumor drug resistance and enhances the efficacy of immunotherapy. Front Oncol. 2023;13. doi:10.3389/fonc.2023.1147239

52. Wurzer H, Hoffmann C, Al Absi A, Thomas C. Actin Cytoskeleton Straddling the Immunological Synapse between Cytotoxic Lymphocytes and Cancer Cells. Cells. 2019;8(5):463. doi:10.3390/cells8050463

53. Wei K, Pan C, Yao G, et al. MiR-106b-5p Promotes Proliferation and Inhibits Apoptosis by Regulating BTG3 in Non-Small Cell Lung Cancer. Cell Physiol Biochem Int J Exp Cell Physiol Biochem Pharmacol. 2017;44(4):1545–1558. doi:10.1159/000485650

54. Hirono T, Jingushi K, Nagata T, et al. MicroRNA-130b functions as an oncomiRNA in non-small cell lung cancer by targeting tissue inhibitor of metalloproteinase-2. Sci Rep. 2019;9(1):6956. doi:10.1038/s41598-019-43355-8

55. Liu Y, Hu X, Hu L, Xu C, Liang X. Let-7i-5p enhances cell proliferation, migration and invasion of ccRCC by targeting HABP4. BMC Urol. 2021;21(1):49. doi:10.1186/s12894-021-00820-9

56. Petersen DL, Berthelsen J, Willerslev-Olsen A, et al. A novel BLK-induced tumor model. Tumor Biol. 2017;39(7):1010428317714196. doi:10.1177/1010428317714196

57. Ishizuka JJ, Manguso RT, Cheruiyot CK, et al. Loss of ADAR1 in tumours overcomes resistance to immune checkpoint blockade. Nature. 2019;565(7737):43-48. doi:10.1038/s41586-018-0768-9

58. Yang X, Li Q, Zeng T. Peripheral CD4+ T cells correlate with response and survival in patients with advanced non-small cell lung cancer receiving chemo-immunotherapy. Front Immunol. 2024;15. doi:10.3389/fimmu.2024.1364507

59. Lambert SL, Zhang C, Guo C, et al. Association of Baseline and Pharmacodynamic Biomarkers With Outcomes in Patients Treated With the PD-1 Inhibitor Budigalimab. J Immunother. 2022;45(3):167. doi:10.1097/CJI.0000000000000408

60. Miao K, Zhang X, Wang H, et al. Peripheral Blood Lymphocyte Subsets Predict the Efficacy of Immune Checkpoint Inhibitors in Non–Small Cell Lung Cancer. Front Immunol. 2022;13. doi:10.3389/fimmu.2022.912180

61. Li P, Qin P, Fu X, et al. Associations between peripheral blood lymphocyte subsets and clinical outcomes in patients with lung cancer treated with immune checkpoint inhibitor. Ann Palliat Med. 2021;10(3):3039–3049. doi:10.21037/apm-21-163

62. Khanniche A, Yang Y, Zhang J, et al. Early-like differentiation status of systemic PD-1+CD8+ T cells predicts PD-1 blockade outcome in non-small cell lung cancer. Clin Transl Immunol. 2022;11(7):e1406. doi:10.1002/cti2.1406

