## Supplementary Material for "Integrative Multi-Omic Profiling of cfDNA Methylation and EV-miRNAs Identifies Immunotherapy-Outcome Molecular Subtypes in NSCLC"

Supplementary Table S1. Univariate Cox regression analysis of markers at baseline (T1) associated with OS or PFS.

Supplementary Table S2. Univariate Cox regression analysis of markers at early treatment (T2) associated with OS or PFS.

Supplementary Table S3. MOFA2 feature loadings for molecular variables in T1 and T2 models.

Supplementary Table S4. Differential expression and methylation analysis across MDC-T1 and MDC-T2 molecular subtypes for each omic layer.

Supplementary Table S5. MDCs immune profiling by flow cytometry.

Supplementary Table S6. Predictive modeling performance for MDC label assignment in external validation cohorts.

Supplementary Figure S1. Sample collection timeline and study workflow overview.

Supplementary Figure S2. Sample availability across time points and omics layers.

Supplementary Figure S3. Optimization of the number of MOFA2 latent factors and clustering resolution.

Supplementary Figure S4. Longitudinal projection of MDCs stratifies NSCLC patients in PFS.

Supplementary Figure S5. Molecular signature heatmaps of differentially expressed miRNAs and methylated regions in MDC subtypes.

Supplementary Figure S6. Clinical associations with MOFA-derived factors and MDC subtypes in T1 and T2 models.

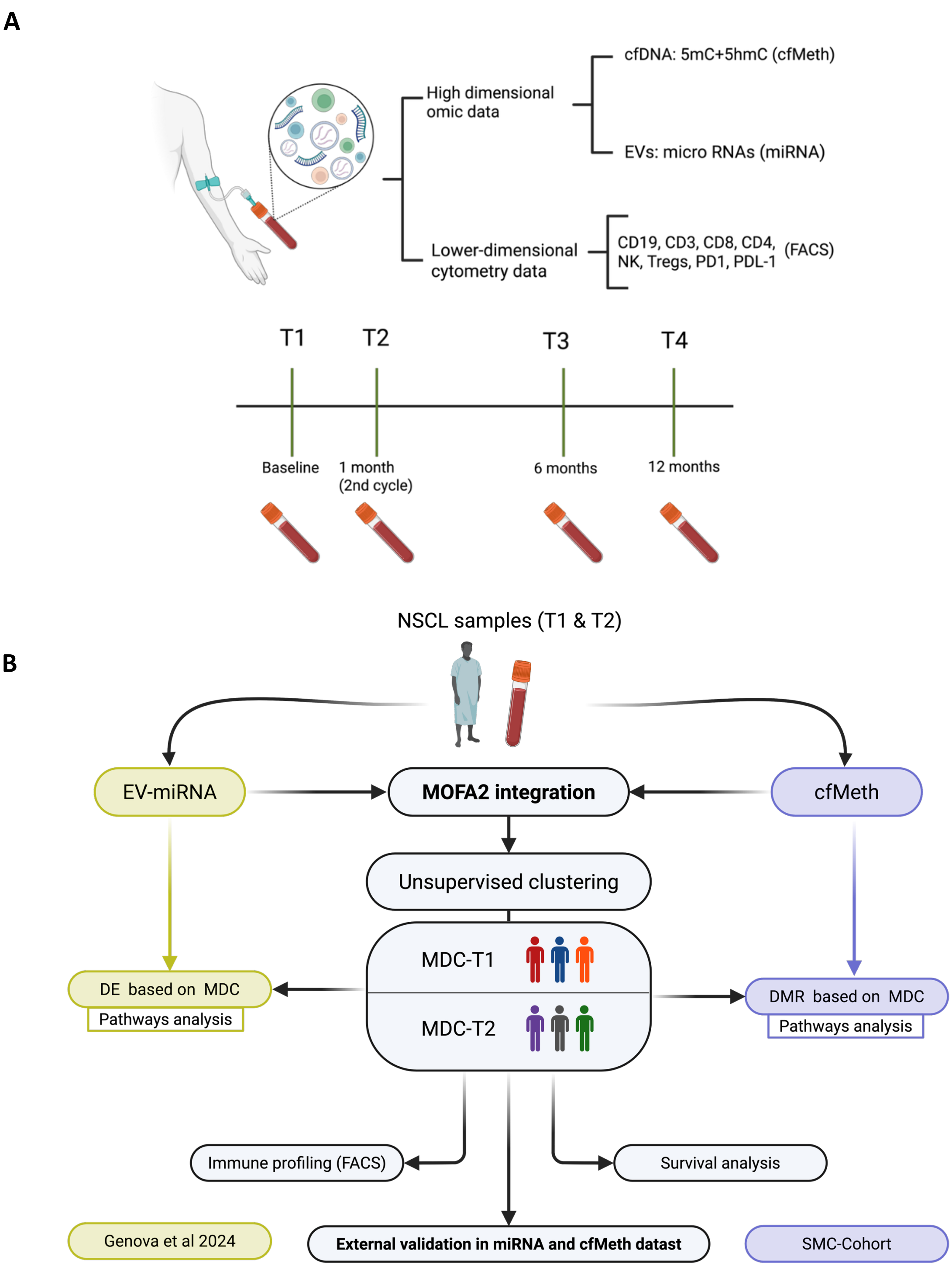


**Supplementary Figure S1. Sample collection timeline and study workflow overview.** (A) Schematic representation of liquid biopsy collection across four predefined time points: T1 (pre-treatment baseline), T2 (after the second cycle of ICI), T3 (month 6), and T4 (month 12). Peripheral blood was collected at each time point to isolate plasma for cfDNA and EV-derived miRNA analyses. (B) Summary of the multi-omic analytical pipeline.

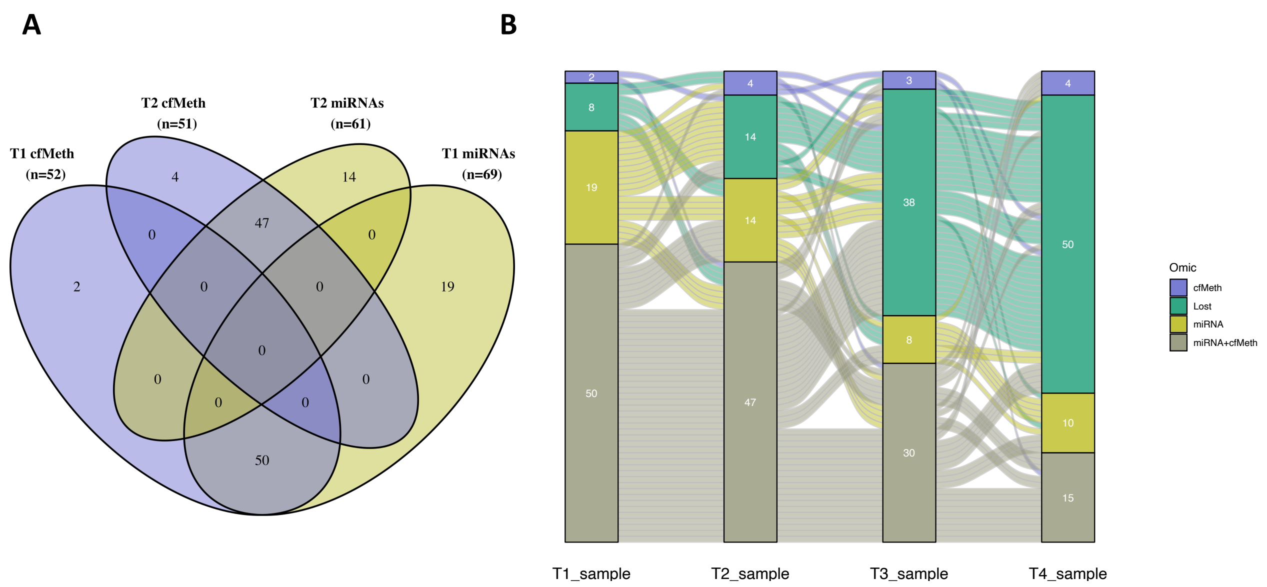


**Supplementary Figure S2. Sample availability across time points and omics layers.** (A) Venn diagram illustrating the distribution of available samples at T1 and T2 for each omic layer: cfDNA methylation (cfMeth) and extracellular vesicle-derived miRNAs. (B) Sankey plot showing the longitudinal sampling trajectory per patient across all four time points (T1-T4), and for each omics modality. The diagram highlights the continuity or dropout of sample availability, as well as single-omic versus multi-omic coverage.

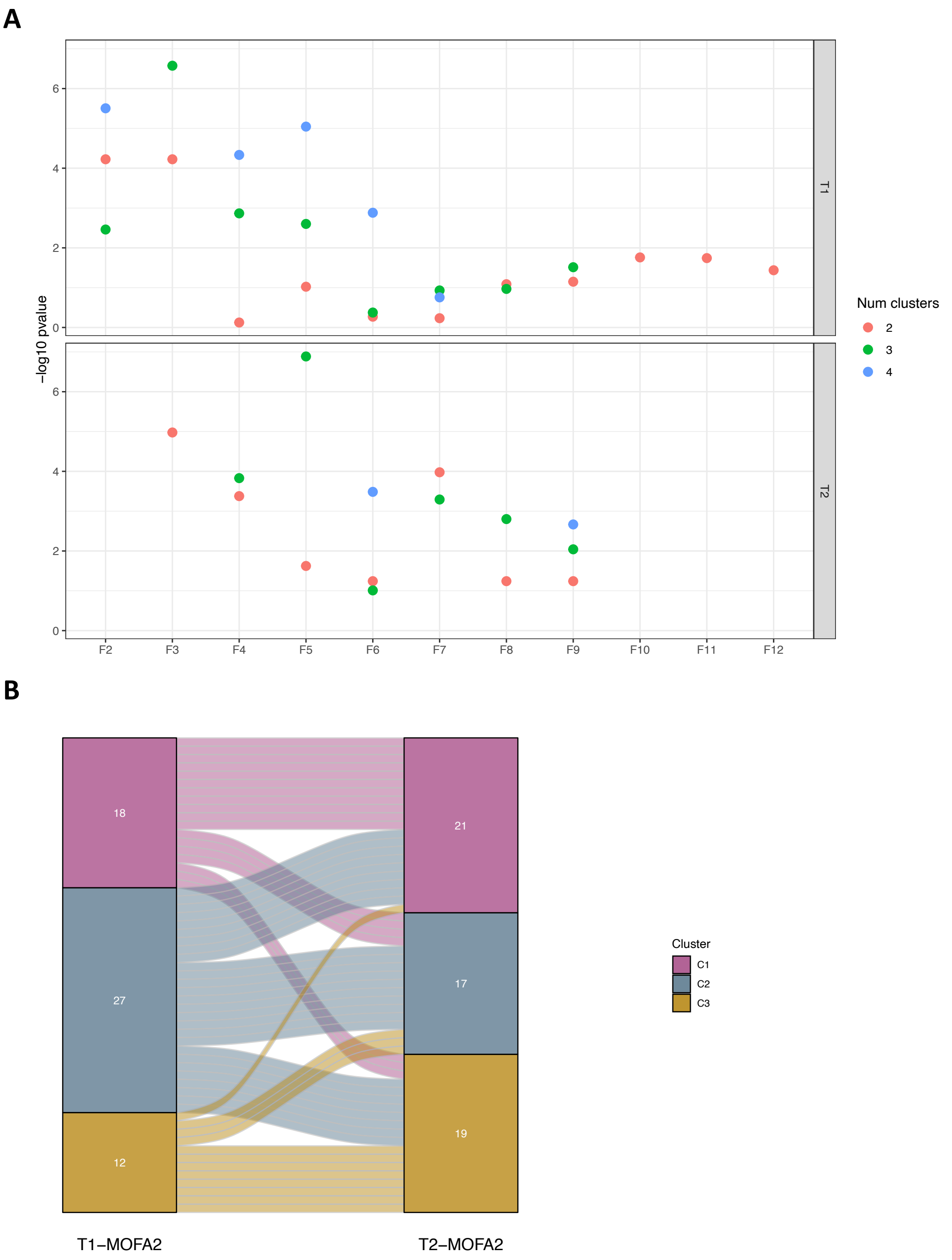


**Supplementary Figure S3. Optimization of the number of MOFA2 latent factors and clustering resolution.** (A) For each time point (T1 and T2), model optimization was guided by survival-based performance metrics. The plot shows the –log10(p-value) from log-rank tests evaluating overall survival stratification, across a range of MOFA2 models with varying numbers of latent factors (x-axis) and clustering resolutions (color-coded by number of clusters). The optimal configuration was determined as the model achieving the most significant survival separation with minimal latent complexity: 3 factors and 3 clusters for T1, and 5 factors and 3 clusters for T2. (B) Sankey diagram illustrating the correspondence between clustering assignments across time points. Each patient's cluster label (based on T1 samples) is connected to their assigned cluster label (based on T2 samples).

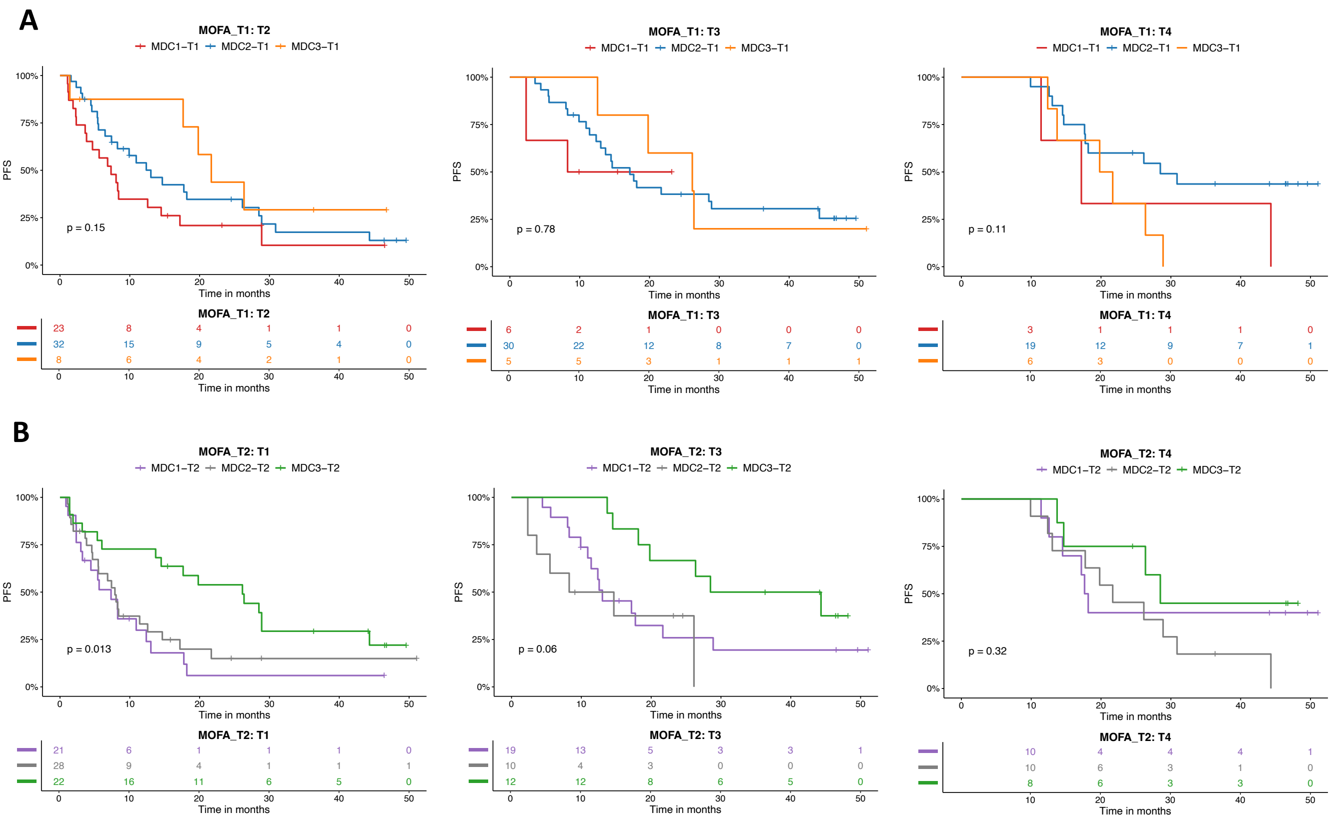


**Supplementary Figure S4. Longitudinal projection of MDCs stratifies NSCLC patients in PFS.** (A) Kaplan–Meier curves for PFS of MDC-T1 subtypes projected onto T2, T3, and T4 samples. (B) Kaplan–Meier curves for PFS in MDC-T2 subtypes projected onto T1, T3, and T4 samples. Subtype assignments were derived from the original MOFA2 models and projected onto new time points using loading matrices. PFS differences were assessed using the log-rank test.

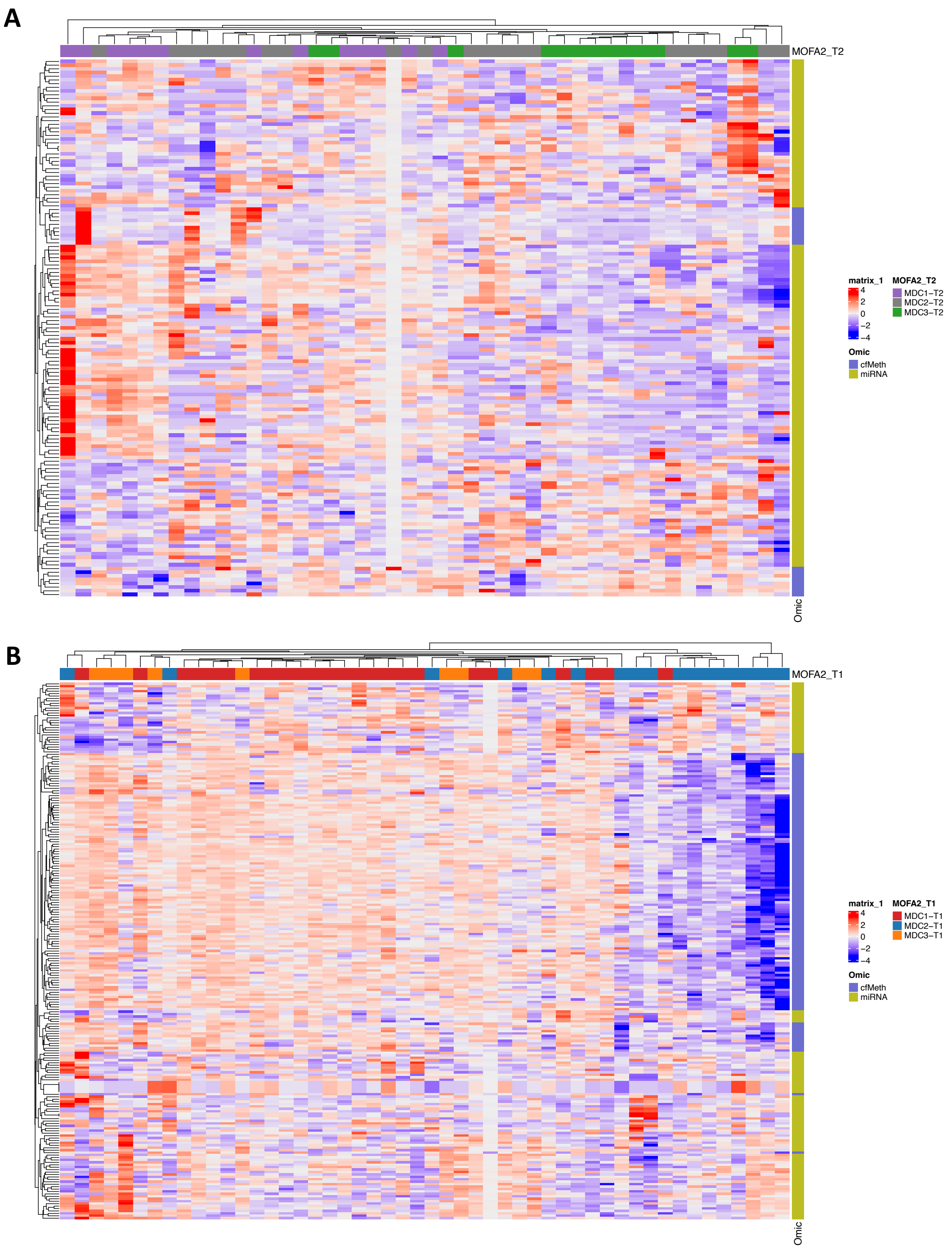


**Supplementary Figure S5. Molecular signature heatmaps of differentially expressed miRNAs and methylated regions in MDC subtypes.** (A) Heatmap displaying differentially expressed miRNAs and differentially methylated regions (DMRs) across MDC-T1 subtypes. (B) Equivalent heatmap for MDC-T2 subtypes. Columns represent individual patient samples annotated by MDC subtype, while rows correspond to features from each omic layer (miRNAs and cfMeth). Hierarchical clustering was applied to the combined, scaled dataset.

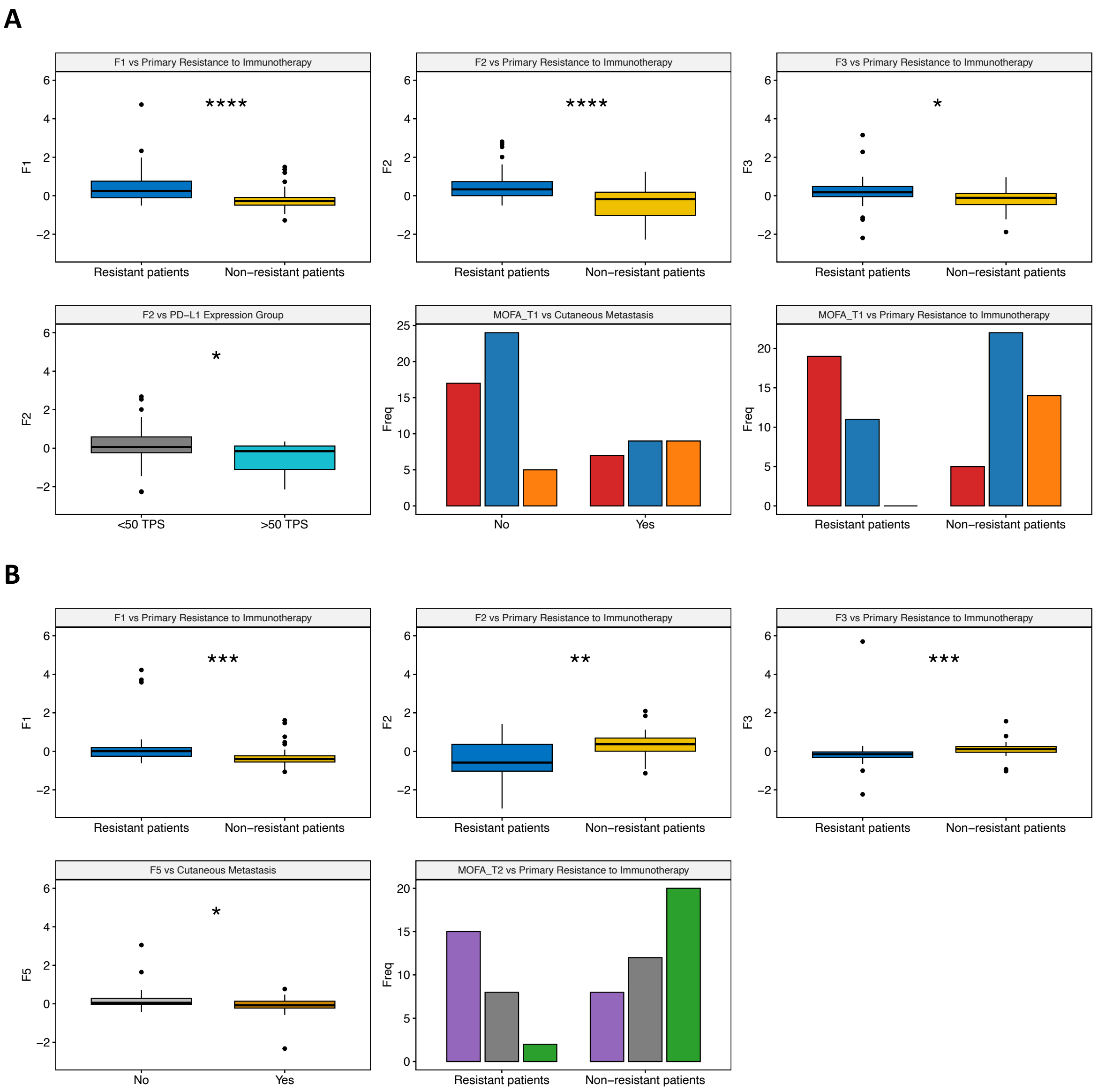


**Supplementary Figure S6. Clinical associations with MOFA-derived factors and MDC subtypes in T1 and T2 models.** (A) MOFA model T1. (B) MOFA model T2. Boxplots represent the distribution of MOFA latent factor values across clinical categories, assessing associations between continuous molecular dimensions and clinical variables. Barplots display contingency tables comparing clinical group distributions across MDC subtypes. Only statistically significant associations are shown. Significance levels are annotated as follows: *** pvalue < 0.001, ** p-value < 0.01, * pvalue < 0.05.
